# Association of Convalescent Plasma Treatment with Reduced Mortality and Improved Clinical Trajectory in Patients Hospitalized with COVID-19 in the Community Setting

**DOI:** 10.1101/2021.06.02.21258190

**Authors:** Shanna A. Arnold Egloff, Angela Junglen, Joseph S.A. Restivo, Marjorie Wongskhaluang, Casey Martin, Pratik Doshi, Daniel Schlauch, Gregg Fromell, Lindsay E. Sears, Mick Correll, Howard A. Burris, Charles F. LeMaistre

## Abstract

**Background:** Convalescent plasma (CP) quickly emerged as one of the first investigational treatment options for COVID-19. Evidence supporting CP for treating patients hospitalized with COVID-19 has been inconclusive, leading to conflicting recommendations regarding its use. The primary objective was to perform a comparative effectiveness study of CP for all-cause, in-hospital mortality in patients with COVID-19.

**Methods:** The matched, multicenter, electronic health records-based, retrospective cohort study included 44,770 patients hospitalized with COVID-19 in one of 176 HCA Healthcare-affiliated community hospitals across the United States from March 2 to October 7, 2020. Coarsened exact matching (1:k) was employed resulting in a sample of 3,774 CP and 10,687 comparison patients.

**Results:** Examining mortality using a shared frailty model and controlling for concomitant medications, calendar date of admission, and days from admission to transfusion demonstrated a significant association of CP with lower risk of mortality compared to the comparison group (aHR = 0.71, 95% CI 0.59-0.86, *p*<0.001). Examination of patient risk trajectories, represented by 400 clinico-demographic features from our Real-Time Risk Model (RTRM), indicated that patients who received CP recovered more quickly. The time from admission to CP transfusion was significantly associated with risk of mortality and stratification revealed that CP within 3 days after admission, but not 4-7 days, was associated with a significant reduction in mortality risk (aHR = 0.53, 95% CI 0.47-0.60, *p*<0.001). CP serology level was inversely associated with mortality when controlling for interaction with days to transfusion (HR = 0.998, 95% CI 0.997-0.999, *p* = 0.013) but was not significant in a univariable analysis.

**Conclusion:** Utilizing this large, diverse, multicenter cohort, we demonstrate that CP is significantly associated with reduced risk of in-hospital mortality. These observations demonstrate the utility of real-world evidence and suggest the need for further evaluation prior to abandoning CP as a viable therapy for COVID-19.

**Funding:** This research was supported, in whole, by HCA Healthcare and/or an HCA Healthcare affiliated entity including Sarah Cannon and Genospace.

**Research in Context:** 

**Evidence before this study:** Discrepant reports of the efficacy of various treatments for COVID-19, including convalescent plasma (CP), emerged from a rapidly evolving political and interventional landscape of the pandemic. Furthermore, clinical interpretations of this discordant data led to underuse, overuse and misuse of certain interventions, often ignoring mechanistic context altogether. CP has been utilized in prior pandemics/epidemics to introduce antibodies to elicit an immune response during the viral phase of infection. Thus, CP received early priority for emergency use and randomized trial engagement. Initially, the United States had issued individual emergency investigational new drug (eIND) use for CP and initiated its expanded access protocol (EAP) to monitor its safety profile and to allow broader access. This effectively restricted access to those with severe disease, which is not mechanistically aligned with targeting the viral phase. Many randomized control trials (RCTs) were being setup for testing efficacy of CP in the inpatient setting and, to a lesser extent, the outpatient setting. Some trial designs focused on severe disease and others on less severe. United States RCTs had additional enrollment challenges due to competing patient access to EAP. All studies were limited by supply and demand due to regional outbreaks and to the shear operational effort of coordinating donations, sampling, serology testing, ordering, and distribution.

To date, most matched studies and RCTs around the globe have shown a trend of CP providing survival benefit, but all had relatively small cohorts except the RECOVERY trial, which failed to show a benefit with CP. Results ranged from no significant effect to 56% reduction in mortality with the latter coming out of a multisite RCT based in New York and Rio De Janeiro. There has been a minimum of nine matched control studies and seven randomized control trials evaluating convalescent plasma.

We frequently assessed World Health Organization (WHO), United Stated Food and Drug Administration (FDA), BARDA/Mayo Clinic led EAP, and the United States Center for Disease Control and Prevention (CDC) resources as well as queried both preprint archives (MedRXIV & SSRN) and PubMed with the search terms “retrospective”, “convalescent plasma”, “randomized”, “trial”, “comparative effect”, “COVID”, “hospital”, “in-hospital”, “hospitalized” and “mortality” to ensure we were considering the most recent methodology and results generated for CP. The last search was performed on May 14, 2021. No date restrictions or language filters were applied.

**Added value of this study:** To our knowledge, this study is the largest and most geographically diverse of its kind to comprehensively evaluate and confirm the beneficial association of CP with all-cause mortality in patients hospitalized with COVID-19. Our data provides context to optimal delivery and validates recent trends in the literature showing CP benefit. There is a dose-response effect with CP antibody levels and we demonstrate that sooner really is better in accordance with the mechanisms of viral clearance and immune regulation. Finally, this is all done in the context of a diverse community setting in one of the largest hospital systems in the United States.

**Implications of all the available evidence:** As novel, more virulent and transmissible SARS-CoV-2 variants emerge around the globe and as reports of post-vaccine “breakthrough” infections and vaccine hesitancy increase, there is a renewed motivation to identify effective treatments for hospitalized patients. The data presented here, along with a growing body of evidence from matched-control studies and RCTs, demonstrate that further evaluation is required prior to abandoning CP as an effective intervention in the treatment of hospitalized COVID-19 patients.

## Introduction

Convalescent plasma (CP) from patients who recover from severe acute respiratory syndrome coronavirus 2 (SARS-CoV-2) has been in use since the inception of the pandemic to treat Coronavirus Disease 2019 (COVID-19) (1-15). As a rapid response to an absence of any FDA approved treatments of this deadly disease, CP use was provided through individual emergency investigational new drug (eIND) and the expanded access protocol (EAP) in the United States, providing sufficient safety data to justify emergency use authorization (EUA) in August of 2020 (16). Although over 100,000 patients were infused under the EAP, the absence of a matched comparison cohort prevents the use of this data to assess the effectiveness of CP (8, 17). Furthermore, randomized control trials (RCTs) struggled with enrollment, design and cohort selection, nimbleness, and aligning site selection with local outbreaks, which resulted in under-powered and/or inconsistent conclusions on the efficacy of convalescent plasma (6, 18-20). Sluggish global deployment of vaccinations, diminished vaccine adoption rates, and the potential appearance of resistant variants have renewed interest in CP. Recent data from the EAP cohort revealed that regional proximity of donor CP to the recipient is associated with reduced mortality, suggesting that regional variations in SARS-CoV-2 could be driving CP responses (21).

Due to the limitations of RCTs in assessing CP during a rapidly evolving pandemic, well-matched, retrospective analyses are critical for comparative effectiveness studies, where they also serve to inform on utilization trends and generate hypotheses. Challenges for retrospective analyses to-date have been the difficulty in accurately generating a matched synthetic control as well as having a sensitive indicator of disease progression and therapeutic response. We previously reported a real-time risk model (RTRM) for COVID-19 that provides a daily granular measure of disease progression to adequately match on baseline disease severity and create a risk trajectory for each patient (22). Using the daily RTRM probabilities and COVID-19 WHO progression scale (WHO PS), we retrospectively examined the association of CP with all-cause, in-hospital mortality and clinical recovery in matched cohorts derived from our COVID-19 registry. The registry consisted of 44,770 patients admitted to one of the 176 HCA Healthcare-affiliated community hospitals where convalescent plasma was provided under an eIND, EAP or EUA.

## Results

### Patient Characteristics

The WHO PS-matched sample resulted in 4,337 CP and 8,708 comparison patients and the RTRM-matched sample resulted in 3,774 CP and 10,687 comparison patients (Figure 1, WHO PS Table S1, RTRM Table 1). The majority of the RTRM-matched sample was Hispanic (49%) or non-Hispanic White (32%), male (60%), and age group 45-64 (46%) with predominant comorbidities of diabetes (30%) and hypertension (49%) (Table 1). For both the CP and comparison groups, 12% of patients presented with severe sepsis and 3% with bacterial pneumonia during their hospitalization (Table 1). Although the difference was minimized by matching, the CP group retained higher rates of sepsis (32%) compared to the comparison group (24%) (Table 1). After excluding those intubated within two days of admission and unlikely to receive CP in time to benefit, there were 1.9% of patients intubated at some point during hospitalization (Table 1). Most patients received anticoagulants, azithromycin, other antibiotics, remdesivir and steroids during hospitalization (Table 1). Biomarker and oxygenation data were descriptively reported for both admission and baseline time points for both CP and comparison groups, which included standard unit transformation and validation of expected values (Table 1).

**Table 1.** Main Model Pre- and Post-RTRM Matching

|  | Pre-Match CP | Pre-Match Comparison | Post-Match CP | Post-Match Comparison |
| --- | --- | --- | --- | --- |
| **Total Number of Patients** | 7393 | 22972 | 3774 | 10687 |
| **Age Groupings<sup>M</sup>** |  |  |  |  |
| 18-44 | 958 (13.3%) | 3970 (17.3%) | 713 (18.9%) | 2019.03 (18.9%) |
| 45-64 | 2861 (38.7%) | 7402 (32.2%) | 1740 (46.1%) | 4927.23 (46.1%) |
| 65-74 | 1707 (23.1%) | 4516 (19.7%) | 667 (17.7%) | 1888.77 (17.7%) |
| 75-84 | 1315 (17.8%) | 4087 (17.8%) | 456 (12.1%) | 1291.28 (12.1%) |
| 85+ | 552 (7.5%) | 2997 (13.0%) | 198 (5.2%) | 560.69 (5.2%) |
| **Race/Ethnicity<sup>M</sup>** |  |  |  |  |
| Hispanic | 3253 (44.0%) | 7500 (32.6%) | 1859 (49.3%) | 5264.21 (49.3%) |
| Non-Hispanic Black | 1150 (15.6%) | 5177 (22.5%) | 512 (13.6%) | 1449.85 (13.6%) |
| Non-Hispanic Other | 460 (6.2%) | 1309 (5.7%) | 215 (5.7%) | 608.82 (5.7%) |
| Non-Hispanic White | 2530 (34.2%) | 8986 (39.1%) | 1188 (31.5%) | 3364.11 (31.5%) |
| **Sex<sup>M</sup>** |  |  |  |  |
| Female | 3082 (41.7%) | 11523 (50.2%) | 1528 (40.5%) | 4326.9 (40.5%) |
| Male | 4311 (58.3%) | 11449 (49.8%) | 2246 (59.5%) | 6360.1 (59.5%) |
| **Smoking Status** |  |  |  |  |
| Current Smoker | 263 (3.6%) | 1322 (5.8%) | 121 (3.2%) | 643.81 (6.0%) |
| Former Smoker | 1444 (19.5%) | 3909 (17.0%) | 609 (16.1%) | 1623.89 (15.2%) |
| Never Smoker | 4815 (65.1%) | 13528 (58.9%) | 2701 (71.6%) | 6838.05 (64.0%) |
| Missing | 871 (11.8%) | 4213 (18.3%) | 343 (9.1%) | 1581.24 (14.8%) |
| **Pre-Admission Comorbidities** |  |  |  |  |
| Asthma or Reactive Airway Disease | 706 (9.5%) | 2315 (10.1%) | 282 (7.5%) | 747.71 (7.0%) |
| COPD (excluding asthma)<sup>M</sup> | 1424 (19.3%) | 4769 (20.8%) | 352 (9.3%) | 996.77 (9.3%) |
| Autoimmune Disorders | 879 (11.9%) | 2826 (12.3%) | 126 (3.3%) | 356.8 (3.3%) |
| Organ-related Autoimmune Disorders<sup>M</sup> | 436 (5.9%) | 1789 (7.8%) | 60 (1.6%) | 169.9 (1.6%) |
| Systemic Autoimmune Disorders<sup>M</sup> | 505 (6.8%) | 1295 (5.6%) | 69 (1.8%) | 195.39 (1.8%) |
| Cancer | 421 (5.7%) | 1442 (6.3%) | 48 (1.3%) | 135.92 (1.3%) |
| Cancer (diagnosed in the last 2 years) | 228 (3.1%) | 795 (3.5%) | 28 (0.7%) | 71.27 (0.7%) |
| Chronic Ischemic Heart Disease<sup>M</sup> | 1544 (20.9%) | 5238 (22.8%) | 368 (9.8%) | 1042.08 (9.8%) |
| Congestive Heart Failure<sup>M</sup> | 1352 (18.3%) | 4648 (20.2%) | 286 (7.6%) | 809.88 (7.6%) |
| Diabetes | 3218 (43.5%) | 9255 (40.3%) | 1144 (30.3%) | 3265.22 (30.6%) |
| Diabetes with Chronic Complications | 1400 (18.9%) | 4327 (18.8%) | 346 (9.2%) | 1005.49 (9.4%) |
| Diabetes without Chronic Complications<sup>M</sup> | 1818 (24.6%) | 4928 (21.5%) | 798 (21.1%) | 2259.73 (21.1%) |
| HIV Infection | 32 (0.4%) | 124 (0.5%) | 10 (0.3%) | 31.22 (0.3%) |
| Hypertension | 4680 (63.3%) | 14966 (65.1%) | 1851 (49%) | 5241.56 (49%) |
| Renal Disease (mild or moderate)<sup>M</sup> | 1147 (15.5%) | 3943 (17.2%) | 298 (7.9%) | 843.86 (7.9%) |
| Renal Disease (severe) | 491 (6.6%) | 1375 (6.0%) | 65 (1.7%) | 184.06 (1.7%) |
| **Medications** |  |  |  |  |
| Anticoagulants | 7232 (97.8%) | 19647 (85.5%) | 3688 (97.7%) | 9519.65 (89.1%) |
| Azithromycin | 5447 (73.7%) | 14123 (61.5%) | 2767 (73.3%) | 6991.49 (65.4%) |
| Antibiotics– Other | 6711 (90.8%) | 18641 (81.1%) | 3278 (86.9%) | 8761.25 (82.0%) |
| Antivirals | 164 (2.2%) | 474 (2.1%) | 54 (1.4%) | 160.58 (1.5%) |
| Hydroxychloroquine | 429 (5.8%) | 2580 (11.2%) | 162 (4.3%) | 331.29 (3.1%) |
| Remdesivir | 4549 (61.5%) | 3162 (13.8%) | 2240 (59.4%) | 2206.96 (20.7%) |
| Tocilizumab | 784 (10.6%) | 554 (2.4%) | 251 (6.7%) | 353.3 (3.3%) |
| Statins/ACEi | 2962 (40.1%) | 8322 (36.2%) | 1209 (32.0%) | 3156.3 (29.5%) |
| Systemic Corticosteroids | 6994 (94.6%) | 12988 (56.5%) | 3563 (94.4%) | 7453.14 (69.7%) |
| Immunomodulators– Other | 11 (0.15%) | 22 (0.1%) | 3 (0.1%) | 5.92 (0.1%) |
| **Clinical Variables** |  |  |  |  |
| Intubation<sup>M</sup> | 1290 (17.4%) | 815 (3.5%) | 71 (1.9%) | 201.05 (1.9%) |
| All Cause Death | 1519 (20.5%) | 1500 (6.5%) | 219 (5.8%) | 477.9 (4.5%) |
| Baseline RTRM Score<sup>M</sup> | 0.14 [0.04, 0.45] | 0.04 [0.02, 0.13] | 0.05 [0.02, 0.12] | 0.03 [0.01, 0.12] |
| **Admission WHO PS** |  |  |  |  |
| WHO PS 2 | 869 (11.8%) | 8019 (34.9%) | 463 (12.3%) | 3427.69 (32.1%) |
| WHO PS 3 | 3816 (51.6%) | 11382 (49.5%) | 2255 (59.8%) | 5316.54 (49.7%) |
| WHO PS 4 | 2524 (34.1%) | 2262 (9.8%) | 1013 (26.8%) | 1365.3 (12.8%) |
| WHO PS 5 | 149 (2.0%) | 283 (1.2%) | 29 (0.8%) | 149.66 (1.4%) |
| Missing | 35 (0.5%) | 1026 (4.5%) | 14 (0.4%) | 427.81 (4.0%) |
| **Baseline WHO PS** |  |  |  |  |
| WHO PS 2 | 247 (3.3%) | 7335 (31.9%) | 175 (4.6%) | 3072.24 (28.7%) |
| WHO PS 3 | 2503 (33.8%) | 11312 (49.2%) | 1827 (48.4%) | 5238.54 (49.0%) |
| WHO PS 4 | 3994 (54.0%) | 2898 (12.6%) | 1701 (45.1%) | 1801.22 (16.9%) |
| WHO PS 5 | 637 (8.6%) | 536 (2.3%) | 61 (1.6%) | 207.07 (1.9%) |
| Missing | 12 (0.2%) | 891 (3.9%) | 10 (0.3%) | 367.93 (3.4%) |
| **Secondary Infections** |  |  |  |  |
| Bacteremia | 44 (0.6%) | 155 (0.6%) | 15 (0.4%) | 66.8 (0.6%) |
| Bacterial Pneumonia | 432 (5.8%) | 682 (3.0%) | 116 (3.1%) | 271.09 (2.5%) |
| Sepsis | 2022 (37.4%) | 5019 (21.8%) | 1202 (31.8%) | 2547.75 (23.8%) |
| Severe Sepsis | 1878 (25.4%) | 2681 (11.7%) | 453 (12.0%) | 1235.86 (11.6%) |
| **Admission Biomarkers** |  |  |  |  |
| Absolute Lymphocyte Count x 10<sup>3</sup>/uL | 0.8 [0.6, 1.2] | 1.0 [0.7, 1.5] | 0.9 [0.7, 1.2] | 1 [0.7, 1.4] |
| Absolute Neutrophil Count x 10<sup>3</sup>/uL | 6.0 [4.0, 8.7] | 5.1 [3.4, 7.6] | 5.9 [4.0, 8.5] | 5.6 [3.8, 8.3] |
| Alanine Aminotransferase units/L | 37 [24, 58] | 31 [21, 51] | 40 [26.5, 63] | 36 [24, 58] |
| Aspartate Aminotransferase units/L | 47 [33, 70] | 38 [26, 59] | 46 [33, 68] | 41 [28, 62] |
| C Reactive Protein mg/dL | 11.2 [6.1, 17.7] | 7.1 [3.2, 13.2] | 10.2 [5.6, 16.9] | 8.1 [3.8, 14.6] |
| D-dimer ng/mL DDU | 510 [315, 880] | 485 [298, 881] | 430 [280, 730] | 446 [280, 750] |
| Ferritin ng/mL | 574 [283, 1066] | 385 [172, 808] | 555 [279, 990] | 461 [216, 916] |
| Hgb A1c | 7.1 [6.2, 9.1] | 6.8 [6.0, 9.0] | 7 [6.1, 9.2] | 7 [6.0, 9.6] |
| Interleukin-6 pg/mL | 46.4 [20.4, 104.0] | 37.2 [14.0, 89.0] | 35.7 [13.9, 84.0] | 37.3 [14.0, 98.0] |
| Lactic Acid Lactate Blood mmol/L | 1.5 [1.2, 2.0] | 1.4 [1.1, 1.9] | 1.5 [1.2, 1.8] | 1.5 [1.1, 1.9] |
| LDH –Serum or Plasma units/L | 379 [286, 512] | 299 [225, 408] | 362 [277, 478] | 318 [239, 442] |
| Procalcitonin ng/mL | 0.23 [0.10, 0.64] | 0.17 [0.09, 0.49] | 0.18 [0.09, 0.43] | 0.17 [0.09, 0.44] |
| Serum Creatinine mg/dL | 1.00 [0.80, 1.40] | 1.0 [0.78, 1.40] | 0.94 [0.77, 1.20] | 0.91 [0.73, 1.20] |
| Total Bilirubin mg/dL | 0.6 [0.4, 0.8] | 0.5 [0.4, 0.8] | 0.6 [0.4, 0.8] | 0.5 [0.4, 0.8] |
| Troponin I ng/mL | 0.03 [0.02, 0.09] | 0.03 [0.02, 0.09] | 0.02 [0.01, 0.06] | 0.03 [0.01, 0.07] |
| White Blood Cell Count x 10<sup>3</sup>/uL | 7.4 [5.4, 10.2] | 7.0 [5.1, 9.7] | 7.3 [5.5, 10.0] | 7.4 [5.4, 10.27] |
| **Baseline Biomarkers** |  |  |  |  |
| Absolute Lymphocyte Count x 10<sup>3</sup>/uL | 0.8 [0.5, 1.2] | 1.1 [0.7, 1.5] | 0.9 [0.6, 1.3] | 1.0 [0.7, 1.5] |
| Absolute Neutrophil Count x 10<sup>3</sup>/uL | 8.1 [5.4, 11.3] | 5.4 [3.5, 8.5] | 7.4 [5.0, 10.2] | 6.21 [3.9, 9.4] |
| Alanine Aminotransferase units/L | 39 [26, 65] | 34 [21, 59] | 44 [28, 72] | 40 [25, 67] |
| Aspartate Aminotransferase units/L | 43 [29, 64] | 37 [25, 59] | 40 [28, 59] | 38 [26, 60] |
| C Reactive Protein mg/dL | 6.6 [3.3, 12.7] | 5.1 [2.3, 9.9] | 5.4 [2.7, 9.8] | 5.0 [2.3, 9.9] |
| D-dimer ng/mL DDU | 565 [326, 1165] | 500 [299, 915] | 426 [269, 782] | 450 [280, 800] |
| Ferritin ng/mL | 606 [328, 1076] | 421 [195, 823] | 555 [297, 939] | 469 [237, 912] |
| Hgb A1c | 6.9 [6.2, 8.6] | 6.9 [6.0, 8.9] | 6.8 [6.1, 8.3] | 7.1 [6.1, 9.4] |
| Interleukin-6 pg/mL. | 32.6 [10.4, 103.1] | 33.3 [10.8, 85.6] | 16.0 [6.7, 62.0] | 29.6 [11.2, 100.9] |
| Lactic Acid Lactate Blood mmol/L | 1.7 [1.3, 2.3] | 1.4 [1.0, 2.0] | 1.5 [1.1, 1.9] | 1.5 [1.1, 2.0] |
| LDH –Serum or Plasma units/L | 401 [297, 556] | 293 [224, 403] | 342 [266, 459] | 301 [232, 411] |
| Procalcitonin ng/mL | 0.24 [0.10, 0.74] | 0.21 [0.09, 0.76] | 0.14 [0.07, 0.34] | 0.16 [0.09, 0.44] |
| Serum Creatinine mg/dL | 0.86 [0.70, 1.20] | 0.87 [0.69, 1.20] | 0.80 [0.66, 1.00] | 0.80 [0.65, 1.02] |
| Total Bilirubin mg/dL | 0.5 [0.4, 0.7] | 0.5 [0.3, 0.7] | 0.5 [0.4, 0.7] | 0.5 [0.3, 0.6] |
| Troponin I ng/mL | 0.10 [0.03, 0.53] | 0.08 [0.03, 0.27] | 0.06 [0.02, 0.40] | 0.07 [0.03, 0.41] |
| White Blood Cell Count x 10<sup>3</sup>/uL | 9.1 [6.5, 12.5] | 7.2 [5.1, 10.1] | 8.6 [6.3, 11.6] | 7.8 [5.4, 10.9] |
| **Admission Oxygenation Measures** |  |  |  |  |
| Arterial Blood Partial Pressure CO2 | 34.2 [30.3, 38.6] | 34.6 [30.2, 39.1] | 34.3 [30.7, 37.8] | 33.87 [30.2, 37.7] |
| Arterial Blood Partial Pressure O2 | 68.6 [59.0, 84.1] | 73.6 [62.1, 93.5] | 69.0 [59.7, 84.4] | 71.8 [61, 90.2] |
| Blood pH | 7.4 [7.4, 7.5] | 7.4 [7.4, 7.5] | 7.4 [7.4, 7.5] | 7.4 [7.4, 7.5] |
| PaO2/FiO2 Ratio | 110.0 [71.0, 207.1] | 234.8 [122.7, 319.5] | 145.6 [83.5, 252.4] | 211.5 [97.7, 305.7] |
| Tidal Volume mL | 480 [433, 550] | 481 [420, 500] | 500 [450, 545] | 500 [450, 550] |
| **Baseline Oxygenation Measures** |  |  |  |  |
| Arterial Blood Partial Pressure CO2 | 37.0 [32.4, 42.6] | 36.0 [31.1, 41.8] | 36.3 [32.4, 40.1] | 35.5 [30.2, 40.2] |
| Arterial Blood Partial Pressure O2 | 69.6 [59.0, 87.5] | 73.0 [61.4, 94.0] | 71.0 [60.5, 86.2] | 73.0 [61.0, 94.0] |
| Blood pH | 7.4 [7.4, 7.5] | 7.4 [7.4, 7.5] | 7.4 [7.4, 7.5] | 7.4 [7.4, 7.5] |
| PaO2/FiO2 Ratio | 89.7 [65.4, 141.3] | 152.0 [87.4, 253.3] | 101.0 [72.5, 164.8] | 123.6 [79.1, 243.0] |
| Tidal Volume mL | 450 [420, 500] | 450 [400, 500] | 450 [421, 500] | 470 [446, 500] |

Categorical data are n (%) and continuous variables are presented as median [IQR]. IQR=interquartile range; RTRM = Real-time Risk Model; CP= Convalescent Plasma; ACEi = angiotensin converting enzyme (ACE) inhibitors; COPD = Chronic obstructive pulmonary disease; WHO PS = World Health Organization Progression Score
<sup>M</sup> indicates a variables used for CEM matching

**Figure 1.**
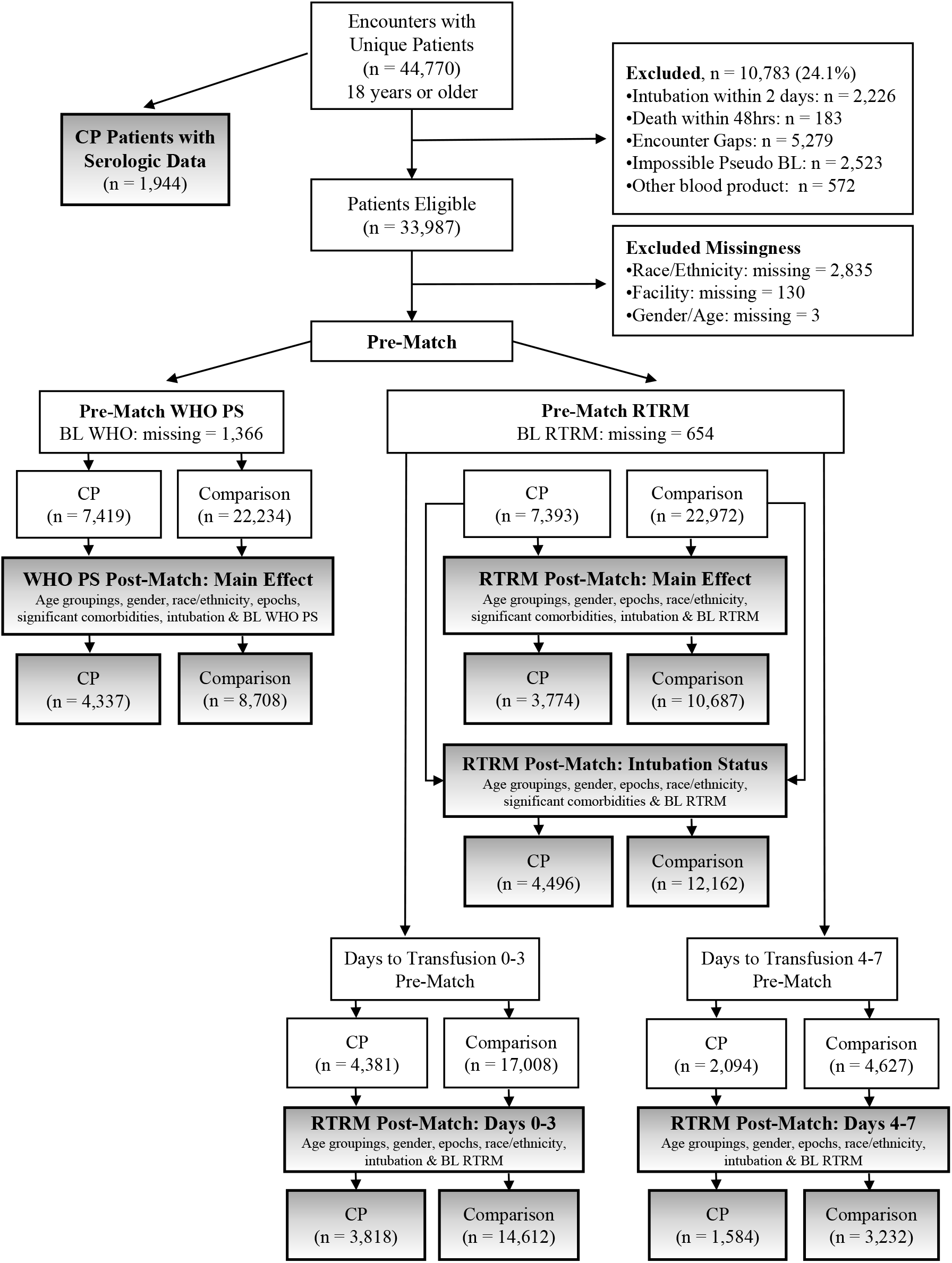
Patient Selection and Matching Processes. Consort diagram displaying patient inclusion/exclusion and filtering criteria for defining the COVID-19 cohort of interest for each analysis. Level of missing data is represented prior to CEM matching. Subsequent pre-match and post-match patient counts are reported with relevant matching criteria that were applied. Grey boxes indicate final post-match cohorts for all reported analyses (WHO PS: Main Effect, RTRM: Main Effect, RTRM: Intubation Status, RTRM: Days to Transfusion 0-3, RTRM: Days to Transfusion 4-7, and Serologic Data).

For the overall CP group (N = 8,034), mean number of days from admission to transfusion and length of stay was 4.0 ± 3.7 and 14.9 ± 10.9, respectively (Table S2). Additional descriptives and frequencies for the overall cohort by calendar epoch can be found in Table S2. Distribution of the frequency of CP transfusions by calendar date is graphed in Figure S1. We evaluated the incidence of serious adverse events related to the CP transfusion and identified a total of 19 (0.2%) events, which included Transfusion-Related Acute Lung Injury (TRALI), Transfusion-Associated Circulatory Overload (TACO), Transfusion Related Infection, Thromboembolic/Thrombotic Event, Severe Allergic Transfusion Reaction, Severe Hemolytic Transfusion Reaction, and Transfusion Related Severe Anaphylaxis.

### All-Cause Mortality

For main analyses examining the effect of CP on all-cause, in-hospital mortality, both the WHO PS-matched and the RTRM-matched models included matching variables of calendar epoch, intubation any time during hospitalization, age grouping, ethnicity/race, sex, significantly different pre-match comorbidities, and severity, measured by the WHO PS score or the RTRM risk probability (using 0.10 increments) at baseline. Significant comorbidities included Organ-Specific Autoimmune Disorder, Systemic Autoimmune Disorder, Chronic Pulmonary Disease (excluding Asthma), Diabetes (without complications), Mild/Moderate Renal Disease, Cancer, Congestive Heart Failure, and Chronic Ischemic Heart Disease (Table 1 and S1).

When examining all-cause, in-hospital mortality using a shared frailty model to account for facility effects, both models, WHO PS-matched and RTRM-matched, demonstrated a significant association of CP with lower risk of mortality compared to the matched comparison group (WHO PS, aHR = 0.75 95% CI 0.65-0.85, *p* < 0.001; RTRM, aHR = 0.71, 95% CI 0.59-0.86, *p* < 0.001) when controlling for concomitant medications, calendar date of admission, and days from admission to transfusion (Table 2). Co-infections such as bacterial pneumonia, sepsis, and severe sepsis were only controlled for in the WHO PS-matched model, since the RTRM accounts for secondary infections.

**Table 2.**
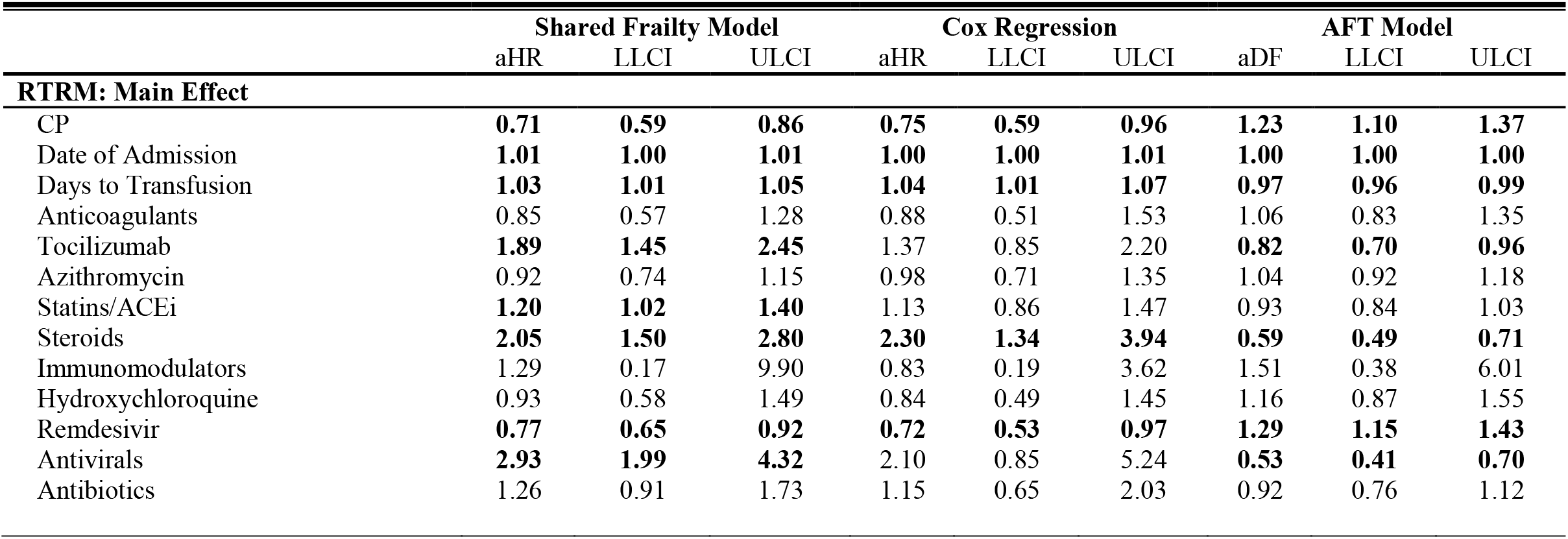
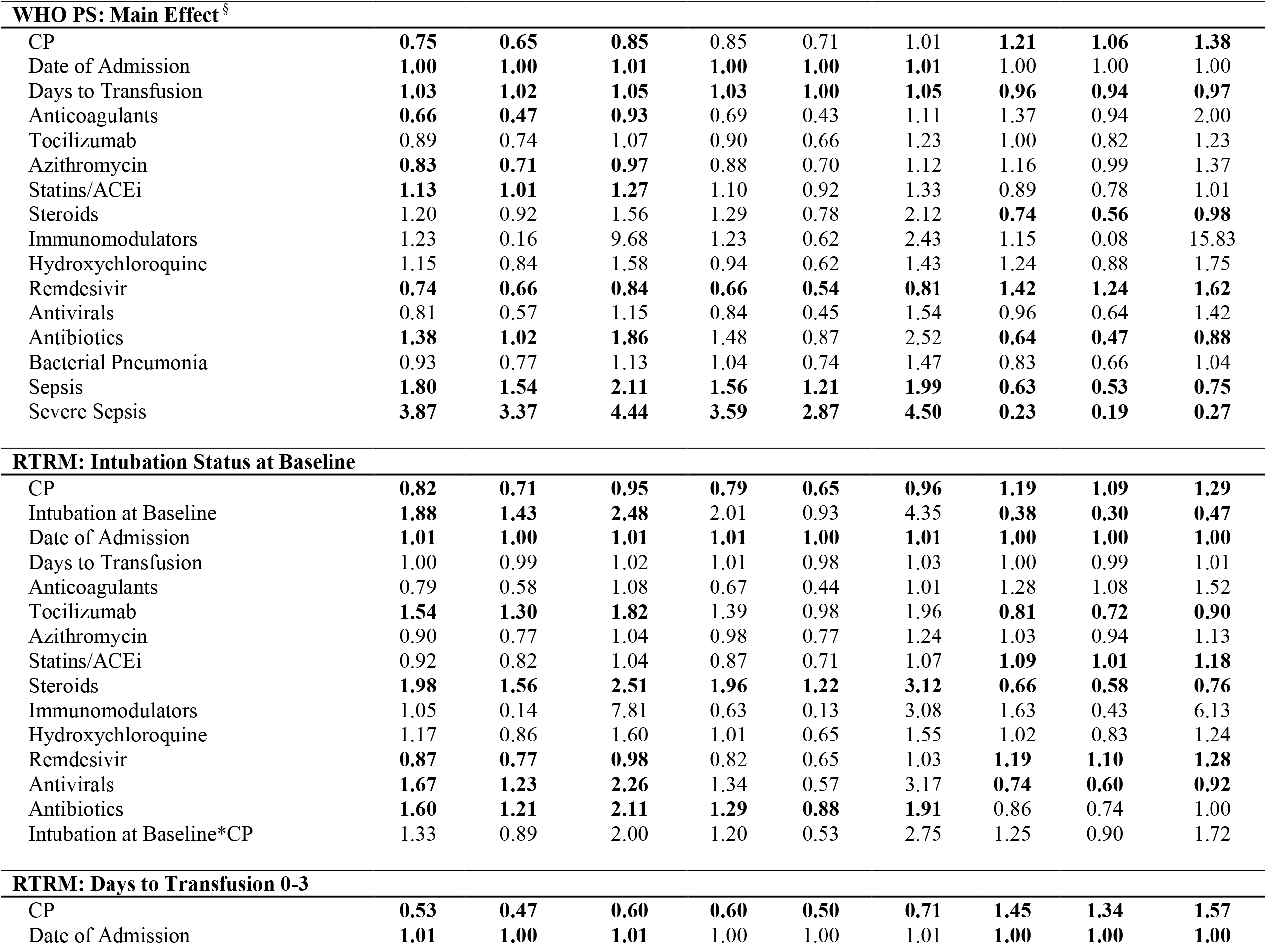
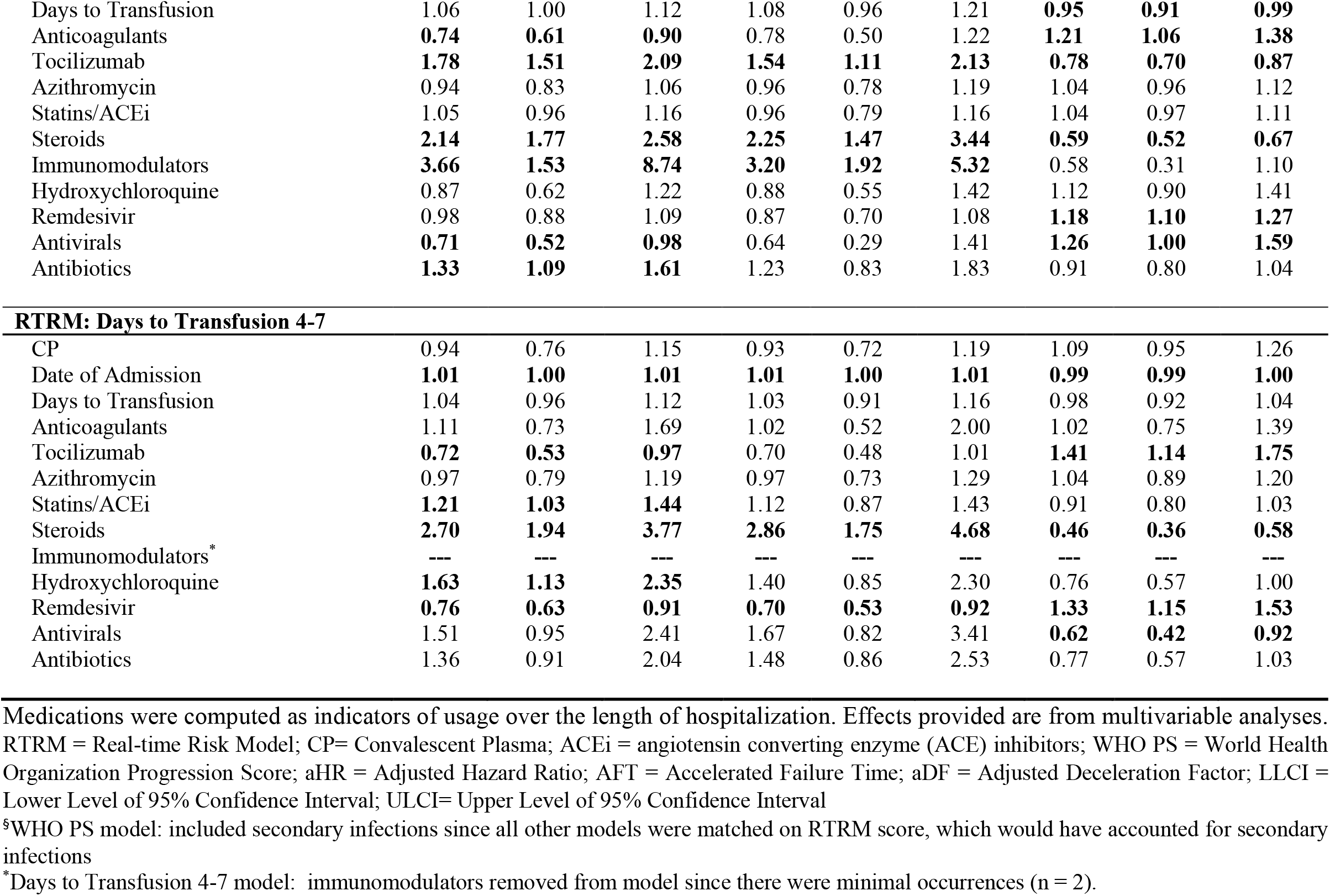
Effects across all Multivariable Mortality Models

|  | Shared Frailty Model |  |  | Cox Regression |  |  | AFT Model |  |  |
| --- | --- | --- | --- | --- | --- | --- | --- | --- | --- |
|  | aHR | LLCI | ULCI | aHR | LLCI | ULCI | aDF | LLCI | ULCI |
| **RTRM: Main Effect** |  |  |  |  |  |  |  |  |  |
| CP | **0.71** | **0.59** | **0.86** | **0.75** | **0.59** | **0.96** | **1.23** | **1.10** | **1.37** |
| Date of Admission | **1.01** | **1.00** | **1.01** | **1.00** | **1.00** | **1.01** | **1.00** | **1.00** | **1.00** |
| Days to Transfusion | **1.03** | **1.01** | **1.05** | **1.04** | **1.01** | **1.07** | **0.97** | **0.96** | **0.99** |
| Anticoagulants | 0.85 | 0.57 | 1.28 | 0.88 | 0.51 | 1.53 | 1.06 | 0.83 | 1.35 |
| Tocilizumab | **1.89** | **1.45** | **2.45** | 1.37 | 0.85 | 2.20 | **0.82** | **0.70** | **0.96** |
| Azithromycin | 0.92 | 0.74 | 1.15 | 0.98 | 0.71 | 1.35 | 1.04 | 0.92 | 1.18 |
| Statins/ACEi | **1.20** | **1.02** | **1.40** | 1.13 | 0.86 | 1.47 | 0.93 | 0.84 | 1.03 |
| Steroids | **2.05** | **1.50** | **2.80** | **2.30** | **1.34** | **3.94** | **0.59** | **0.49** | **0.71** |
| Immunomodulators | 1.29 | 0.17 | 9.90 | 0.83 | 0.19 | 3.62 | 1.51 | 0.38 | 6.01 |
| Hydroxychloroquine | 0.93 | 0.58 | 1.49 | 0.84 | 0.49 | 1.45 | 1.16 | 0.87 | 1.55 |
| Remdesivir | **0.77** | **0.65** | **0.92** | **0.72** | **0.53** | **0.97** | **1.29** | **1.15** | **1.43** |
| Antivirals | **2.93** | **1.99** | **4.32** | 2.10 | 0.85 | 5.24 | **0.53** | **0.41** | **0.70** |
| Antibiotics | 1.26 | 0.91 | 1.73 | 1.15 | 0.65 | 2.03 | 0.92 | 0.76 | 1.12 |

| **WHO PS: Main Effect <sup>§</sup>** |  |  |  |  |  |  |  |  |  |
| --- | --- | --- | --- | --- | --- | --- | --- | --- | --- |
| CP | **0.75** | **0.65** | **0.85** | 0.85 | 0.71 | 1.01 | **1.21** | **1.06** | **1.38** |
| Date of Admission | **1.00** | **1.00** | **1.01** | **1.00** | **1.00** | **1.01** | 1.00 | 1.00 | 1.00 |
| Days to Transfusion | **1.03** | **1.02** | **1.05** | **1.03** | **1.00** | **1.05** | **0.96** | **0.94** | **0.97** |
| Anticoagulants | **0.66** | **0.47** | **0.93** | 0.69 | 0.43 | 1.11 | 1.37 | 0.94 | 2.00 |
| Tocilizumab | 0.89 | 0.74 | 1.07 | 0.90 | 0.66 | 1.23 | 1.00 | 0.82 | 1.23 |
| Azithromycin | **0.83** | **0.71** | **0.97** | 0.88 | 0.70 | 1.12 | 1.16 | 0.99 | 1.37 |
| Statins/ACEi | **1.13** | **1.01** | **1.27** | 1.10 | 0.92 | 1.33 | 0.89 | 0.78 | 1.01 |
| Steroids | 1.20 | 0.92 | 1.56 | 1.29 | 0.78 | 2.12 | **0.74** | **0.56** | **0.98** |
| Immunomodulators | 1.23 | 0.16 | 9.68 | 1.23 | 0.62 | 2.43 | 1.15 | 0.08 | 15.83 |
| Hydroxychloroquine | 1.15 | 0.84 | 1.58 | 0.94 | 0.62 | 1.43 | 1.24 | 0.88 | 1.75 |
| Remdesivir | **0.74** | **0.66** | **0.84** | **0.66** | **0.54** | **0.81** | **1.42** | **1.24** | **1.62** |
| Antivirals | 0.81 | 0.57 | 1.15 | 0.84 | 0.45 | 1.54 | 0.96 | 0.64 | 1.42 |
| Antibiotics | **1.38** | **1.02** | **1.86** | 1.48 | 0.87 | 2.52 | **0.64** | **0.47** | **0.88** |
| Bacterial Pneumonia | 0.93 | 0.77 | 1.13 | 1.04 | 0.74 | 1.47 | 0.83 | 0.66 | 1.04 |
| Sepsis | **1.80** | **1.54** | **2.11** | **1.56** | **1.21** | **1.99** | **0.63** | **0.53** | **0.75** |
| Severe Sepsis | **3.87** | **3.37** | **4.44** | **3.59** | **2.87** | **4.50** | **0.23** | **0.19** | **0.27** |
| **RTRM: Intubation Status at Baseline** |  |  |  |  |  |  |  |  |  |
| CP | **0.82** | **0.71** | **0.95** | **0.79** | **0.65** | **0.96** | **1.19** | **1.09** | **1.29** |
| Intubation at Baseline | **1.88** | **1.43** | **2.48** | 2.01 | 0.93 | 4.35 | **0.38** | **0.30** | **0.47** |
| Date of Admission | **1.01** | **1.00** | **1.01** | **1.01** | **1.00** | **1.01** | **1.00** | **1.00** | **1.00** |
| Days to Transfusion | 1.00 | 0.99 | 1.02 | 1.01 | 0.98 | 1.03 | 1.00 | 0.99 | 1.01 |
| Anticoagulants | 0.79 | 0.58 | 1.08 | 0.67 | 0.44 | 1.01 | 1.28 | 1.08 | 1.52 |
| Tocilizumab | **1.54** | **1.30** | **1.82** | 1.39 | 0.98 | 1.96 | **0.81** | **0.72** | **0.90** |
| Azithromycin | 0.90 | 0.77 | 1.04 | 0.98 | 0.77 | 1.24 | 1.03 | 0.94 | 1.13 |
| Statins/ACEi | 0.92 | 0.82 | 1.04 | 0.87 | 0.71 | 1.07 | **1.09** | **1.01** | **1.18** |
| Steroids | **1.98** | **1.56** | **2.51** | **1.96** | **1.22** | **3.12** | **0.66** | **0.58** | **0.76** |
| Immunomodulators | 1.05 | 0.14 | 7.81 | 0.63 | 0.13 | 3.08 | 1.63 | 0.43 | 6.13 |
| Hydroxychloroquine | 1.17 | 0.86 | 1.60 | 1.01 | 0.65 | 1.55 | 1.02 | 0.83 | 1.24 |
| Remdesivir | **0.87** | **0.77** | **0.98** | 0.82 | 0.65 | 1.03 | **1.19** | **1.10** | **1.28** |
| Antivirals | **1.67** | **1.23** | **2.26** | 1.34 | 0.57 | 3.17 | **0.74** | **0.60** | **0.92** |
| Antibiotics | **1.60** | **1.21** | **2.11** | **1.29** | **0.88** | **1.91** | 0.86 | 0.74 | 1.00 |
| Intubation at Baseline*CP | 1.33 | 0.89 | 2.00 | 1.20 | 0.53 | 2.75 | 1.25 | 0.90 | 1.72 |
| **RTRM: Days to Transfusion 0-3** |  |  |  |  |  |  |  |  |  |
| CP | **0.53** | **0.47** | **0.60** | **0.60** | **0.50** | **0.71** | **1.45** | **1.34** | **1.57** |
| Date of Admission | **1.01** | **1.00** | **1.01** | 1.00 | 1.00 | 1.01 | **1.00** | **1.00** | **1.00** |
| Days to Transfusion | 1.06 | 1.00 | 1.12 | 1.08 | 0.96 | 1.21 | **0.95** | **0.91** | **0.99** |
| Anticoagulants | **0.74** | **0.61** | **0.90** | 0.78 | 0.50 | 1.22 | **1.21** | **1.06** | **1.38** |
| Tocilizumab | **1.78** | **1.51** | **2.09** | **1.54** | **1.11** | **2.13** | **0.78** | **0.70** | **0.87** |
| Azithromycin | 0.94 | 0.83 | 1.06 | 0.96 | 0.78 | 1.19 | 1.04 | 0.96 | 1.12 |
| Statins/ACEi | 1.05 | 0.96 | 1.16 | 0.96 | 0.79 | 1.16 | 1.04 | 0.97 | 1.11 |
| Steroids | **2.14** | **1.77** | **2.58** | **2.25** | **1.47** | **3.44** | **0.59** | **0.52** | **0.67** |
| Immunomodulators | **3.66** | **1.53** | **8.74** | **3.20** | **1.92** | **5.32** | 0.58 | 0.31 | 1.10 |
| Hydroxychloroquine | 0.87 | 0.62 | 1.22 | 0.88 | 0.55 | 1.42 | 1.12 | 0.90 | 1.41 |
| Remdesivir | 0.98 | 0.88 | 1.09 | 0.87 | 0.70 | 1.08 | **1.18** | **1.10** | **1.27** |
| Antivirals | **0.71** | **0.52** | **0.98** | 0.64 | 0.29 | 1.41 | **1.26** | **1.00** | **1.59** |
| Antibiotics | **1.33** | **1.09** | **1.61** | 1.23 | 0.83 | 1.83 | 0.91 | 0.80 | 1.04 |
| RTRM: Days to Transfusion 4-7 |  |  |  |  |  |  |  |  |  |
| CP | 0.94 | 0.76 | 1.15 | 0.93 | 0.72 | 1.19 | 1.09 | 0.95 | 1.26 |
| Date of Admission | **1.01** | **1.00** | **1.01** | **1.01** | **1.00** | **1.01** | **0.99** | **0.99** | **1.00** |
| Days to Transfusion | 1.04 | 0.96 | 1.12 | 1.03 | 0.91 | 1.16 | 0.98 | 0.92 | 1.04 |
| Anticoagulants | 1.11 | 0.73 | 1.69 | 1.02 | 0.52 | 2.00 | 1.02 | 0.75 | 1.39 |
| Tocilizumab | **0.72** | **0.53** | **0.97** | 0.70 | 0.48 | 1.01 | **1.41** | **1.14** | **1.75** |
| Azithromycin | 0.97 | 0.79 | 1.19 | 0.97 | 0.73 | 1.29 | 1.04 | 0.89 | 1.20 |
| Statins/ACEi | **1.21** | **1.03** | **1.44** | 1.12 | 0.87 | 1.43 | 0.91 | 0.80 | 1.03 |
| Steroids | **2.70** | **1.94** | **3.77** | **2.86** | **1.75** | **4.68** | **0.46** | **0.36** | **0.58** |
| Immunomodulators* | --- | --- | --- | --- | --- | --- | --- | --- | --- |
| Hydroxychloroquine | **1.63** | **1.13** | **2.35** | 1.40 | 0.85 | 2.30 | 0.76 | 0.57 | 1.00 |
| Remdesivir | **0.76** | **0.63** | **0.91** | **0.70** | **0.53** | **0.92** | **1.33** | **1.15** | **1.53** |
| Antivirals | 1.51 | 0.95 | 2.41 | 1.67 | 0.82 | 3.41 | **0.62** | **0.42** | **0.92** |
| Antibiotics | 1.36 | 0.91 | 2.04 | 1.48 | 0.86 | 2.53 | 0.77 | 0.57 | 1.03 |

Medications were computed as indicators of usage over the length of hospitalization. Effects provided are from multivariable analyses. RTRM = Real-time Risk Model; CP= Convalescent Plasma; ACEi = angiotensin converting enzyme (ACE) inhibitors; WHO PS = World Health Organization Progression Score; aHR = Adjusted Hazard Ratio; AFT = Accelerated Failure Time; aDF = Adjusted Deceleration Factor; LLCI = Lower Level of 95% Confidence Interval; ULCI= Upper Level of 95% Confidence Interval
<sup>§</sup>WHO PS model: included secondary infections since all other models were matched on RTRM score, which would have accounted for secondary infections
\*Days to Transfusion 4-7 model: immunomodulators removed from model since there were minimal occurrences (n = 2).

Both the WHO PS-matched model and RTRM-matched model violated the assumption of proportional hazards, so Accelerated Failure Time (AFT) models were performed to examine the consistency of the effect (Table 2). The AFT did replicate the main findings of CP on life expectancy for both models but the AFT models did not account for facility effects (WHO PS, aDF = 1.21, 95% CI 1.06, 1.38, *p* = 0.005; RTRM, aDF = 1.23, 95% CI 1.10, 1.37, *p* = 0.005).

### RTRM Risk Trajectories

We investigated the effect of CP on rate of recovery using a mixed effects model with the RTRM-matched sample to evaluate RTRM trajectories over time and account for facility effects. CP was significantly associated with a quicker RTRM score improvement than the matched comparison group, controlling for concomitant medications, calendar date of admission, and days from admission to transfusion (*p* < 0.001). Patients treated with CP were associated with a quicker decline in risk severity over time than the matched comparison group for both overall hospitalization (CP, *b* = -0.0038; Comparison, *b* = -0.0030) and the first 10-day window (CP, *b* = -0.0040; Comparison, *b* = -0.0034), respectively. This -0.0008 difference in slope equated to an overall 27% difference in risk reduction per day of hospitalization for CP relative to the comparison group. The RTRM risk trajectory analyses assumed a linear fit, so we provided average daily RTRM probabilities over hospitalization to observe non-linear trends in disease progression for the RTRM Main Effect, Intubation Status, and Days to Transfusion (Figure 2).

**Figure 2:**
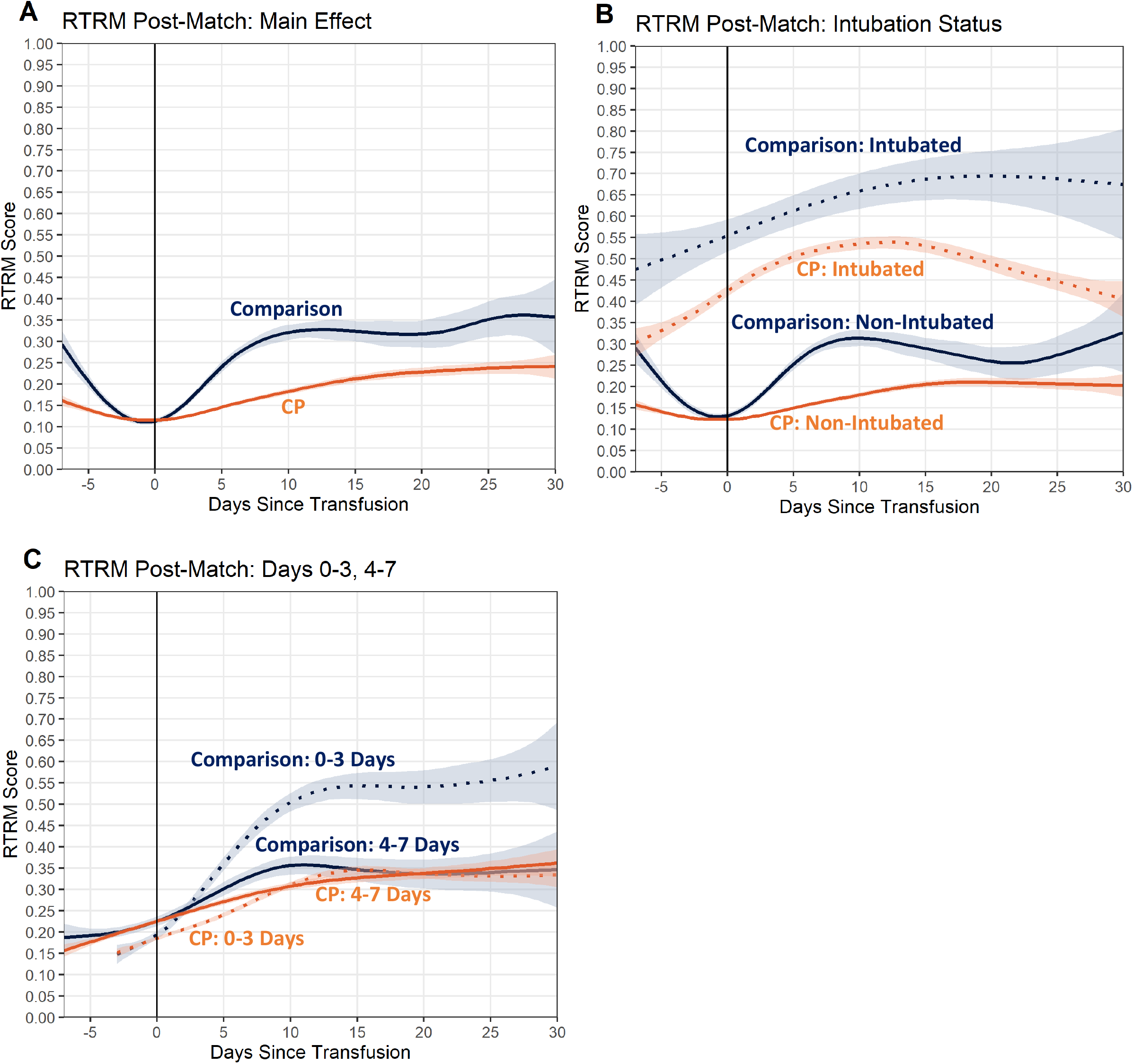
Real-Time Risk Curves. RTRM curves, smoothed and weighted, showing risk trajectories across CP and the comparison group. A generalized additive model with integrated smoothness estimation was applied to the risk predictions over hospitalization time, which were anchored by baseline date. Patients at discharge were assigned a final RTRM probability based on vital status of 1.00 for expired and 0.00 for alive. Shaded boundaries around each curve represent the 95% confidence intervals (95%). A) Depicts the main weighted comparison of the RTRM-matched cohort of CP (orange) and the comparison (blue) groups. B) Includes the weighted comparison of CP (orange) and the comparison (blue) groups stratified by intubation (dotted) versus no intubation (solid) at any point during hospitalization. C) Compilation of weighted comparisons between CP (orange) and the comparison (blue) groups for each of the two 0-3 (dotted) and 4-7 (solid) days from admission to transfusion groupings.

### Intubation Status Subgroups

Intubation status was defined by whether patients were intubated prior to or on same day as transfusion. Statistical analyses examined the interaction of CP with intubation, which serves as a surrogate for baseline severity, in relation to all-cause, in-hospital mortality. The shared frailty analysis showed that the effect of CP on risk of mortality did not differ for patients intubated prior to or at baseline compared to non-intubated patients, as demonstrated by including an interaction term in the model and controlling for concomitant medications, calendar date, and days from admission to transfusion (*p* = 0.160; Table 2).

### Days to Transfusion

There was a significant effect of days from admission to transfusion on mortality, specifically within the CP group (aHR = 1.06, 95% CI 1.03-1.09, *p* < 0.001). Therefore, exploratory analyses were performed to detail this effect. A Cox regression model was progressively run, splitting by every number of days from admission to transfusion, and it was noted that the upper 95% confidence interval of the aHR did not cross 1.0 out to 7 days. However, if each number of days was run in isolation, 3 days from admission was the limit due to decreasing sample size.

Therefore, we examined the effect of CP across days on mortality risk by stratifying the RTRM sample into two groups, 0-3 and 4-7 days, in accordance with sample distribution. Each CP group was matched to comparison on baseline RTRM probability, sex, age and race. We identified a significant association of CP with reduced mortality risk when examining the 0-3 day group (aHR = 0.53, 95% CI 0.47-0.60, *p* < 0.001), but this association was not significant for the 4-7 day group (aHR = 0.94, 95% CI 0.76-1.15, *p* = 0.520) after adjusting for concomitant medications, calendar date, and days from admission to transfusion, suggesting the significant association of days from admission to transfusion is driven by the first 3 days. However, this should not be interpreted as there is no benefit after 3 days with CP, as the distribution of our sample was dominated by CP transfusions within 3 days after admission (66%, Figure S2).

### Serology

When examining total anti-SARS-CoV-2 S/Co serology as either a continuous variable or as ordinal low, medium, high (20^th^ and 80^th^ percentiles), there was no significant association with risk of mortality, albeit there was a trend in the expected direction (n=1,944, Table S3) (8). To further this exploration, we examined whether the impact of serologic levels on mortality was influenced by days from admission to transfusion. The interaction of S/Co serology level as a continuous variable with days to transfusion was significantly associated with risk of mortality (*p* = 0.044), along with the main effects (Serology, HR = 0.998, 95% CI 0.997-0.999, *p* = 0.013; Days to Transfusion, HR = 1.036, 95% CI 1.002-1.071, *p* = 0.037) (Table S3). This resulted in a 0.2% decreased risk of mortality for every 1 unit of S/Co serology level, where it ranged from 1.25 to 932.00 with a mean of 178.21 and standard deviation of 138.49. Simple slopes analyses were performed to examine this significant interaction and reported in Table S3.

## Discussion

Although others have shown a correlation with the levels of antibody titers in CP with improved outcomes and there are clear trends towards benefit across the COVID-19 CP literature,(8, 23) our data is the first to provide definitive evidence in a nationwide, community-based matched cohort that CP is associated with a 29% reduced risk of death in patients hospitalized with COVID-19. This effect is even more pronounced if CP is delivered within the first three days after admission, revealing a 47% reduction in risk of in-hospital mortality for all patients regardless of baseline severity. However, detailed analyses suggested there might be continued benefit beyond day 3 indicating further investigations are warranted. Moreover, patients treated with CP experienced a faster recovery equating to a 27% difference in reduction in risk/severity per day over their length of stay as measured by daily RTRM probabilities. Interestingly, this effect was not dependent on baseline severity, as we found no differential association of CP with mortality risk based on intubated status prior to or at baseline.

SARS-CoV-2 antibody levels within a subset of CP patients, as measured by total S/Co, were associated with a 0.2% reduced risk of mortality per unit increase after adjusting for its interaction with days from admission to transfusion. Joyner *et al*. have also shown within the large EAP cohort that semi-quantitative CP antibody levels were inversely correlated with mortality when given within a few days of hospitalization (8). Despite methodological differences in laboratory approaches and serology assay platforms used in our two studies, the consistency across these studies suggest there is, indeed, a dose-response benefit to receiving CP.

Although we made substantial efforts to manage challenges of analyzing real-world data, there are limitations inherent in their use. These include the evolution of diagnosis and treatment during this pandemic as well as changes in medical documentation and coding related to COVID-19 across multiple facilities. We attempted to account for this by matching on calendar epochs, controlling for calendar date, and nesting on facility. Neither BMI nor smoking status could be included in our analyses due to unreliability and missingness, respectively. Incorporating days from symptom onset to transfusion, rather than days from admission, could more accurately identify optimal timing of CP transfusion. However, less than 10% of patients had clear reporting of symptom onset and, thus, we could not confidently execute analyses with symptom onset due to missingness. In addition, although concomitant medications were controlled for in all primary, secondary and subgroup analyses, they were treated as indicators during hospitalization rather than accounting for their timing with CP and dose. Future studies will be directed at evaluating medication interactions with CP and identifying optimal dosing and timing of concomitant medications.

A challenge to retrospectively evaluating CP delivered in the community setting has been the evolution of treatment criteria and access to CP. The CP cohort was skewed to higher severity based on eligibility criteria for EAP enrollment criteria prior to approval for EUA. Inconsistencies in supply-and-demand occurred throughout 2020 in various outbreak locations. These challenges made it difficult for most research groups to create a well-matched comparison cohort; however, we were able to capitalize on the granularity of our RTRM and the coarsened exact matching approach to match on hundreds of clinico-demographic and biomarker features to identify a properly matched cohort (22). Although RCTs are the gold-standard for assessing efficacy, their deployment during a rapidly evolving pandemic is especially challenging at-scale and in the community setting. Our robust comparative effectiveness evaluation of CP utilizing the large and diverse HCA Healthcare COVID-19 registry for rigorous matching demonstrates a significant reduction in mortality in hospitalized patients, especially those treated within 3 days of admission. This retrospective study gets as close to mitigating the biases arising from non-randomized treatment assignment as one can achieve. Indeed, our data aligns with a recently published multisite RCT, which demonstrates a 56% reduction in 28-day in-hospital mortality (20). Finally, these results corroborate work from Casadevall and colleagues showing an inverse correlation (−0.52) between the number of COVID-19 deaths occurring within two weeks from hospital admission and CP usage within the Unites States (24). We believe our data to be applicable to diverse groups as well as important in contemplating design of future RCTs.

As novel, more virulent and transmissible SARS-CoV-2 variants emerge around the globe and as reports of post-vaccine “breakthrough” infections and vaccine hesitancy increase, there is a renewed motivation to identify effective treatments for hospitalized patients. The data presented here demonstrate that further evaluation is required prior to abandoning CP as an effective intervention in the treatment of hospitalized COVID-19 patients.

## Methods

### Data Sources

The design, analysis, and data interpretations were conducted independently by the investigators. All authors testify to the accuracy and completeness of the data with acknowledgement that there are limitations to real-world evidence. Data for the study was obtained through HCA Healthcare’s data warehouse, which contains detailed and structured clinico-demographic, medical, medical history, pharmacy, laboratory, and outcomes data captured in electronic medical record systems (Epic, Cerner, Meditech) of 176 HCA Healthcare-affiliated clinics and community hospitals across the United States. Any data captured from March 2 to October 7, 2020 were included in the study, allowing for a minimum of 10-day follow-up, while still retaining 99% of the initial sample with available discharge dates (Figure 1).

### Patient Selection

Patients, 18 years or older, that were hospitalized at any of HCA Healthcare’s hospitals and tested positive (CDC confirmed) or presumptive positive (not CDC confirmed) for SARS-CoV-2 by any assay platform (PCR, Rapid Antigen, Antibody etc.) four weeks before to two weeks after their admission date were eligible for the study. Patients were excluded if they were intubated or expired within the first 48 hours after admission. Any data from prior or subsequent admissions or transfers within 36 hours were linked to create a continuum of patient care (Supplemental Materials, Patient Encounters). Patients were excluded if there were discordant gaps in care across data sources (Figure 1).

### Exposure

The primary exposure for comparison was the first transfusion of convalescent plasma (CP) for treatment of COVID-19. We identified CP exposure by patients having a transfusion date, receiving a blood product with a variation of convalescent and/or COVID in the name, and a corresponding ISBT-128 CP barcode as confirmed by blood-bank suppliers. Patients who received unconfirmed plasma products were excluded from analyses. Patients were included regardless of whether CP access was provided under an eIND, EAP, or EUA. Patients that did not receive transfusion of CP or any other plasma product were considered as the comparison group.

We were able to obtain Ortho Vitros serology signal to cut-off ratio (S/Co) data measuring total anti-SARS-CoV-2 antibodies from blood-bank suppliers for a subset of patients treated with CP. Positive serology was defined as a S/Co>1 (n=1944). Mean serology levels were calculated for patients receiving multiple donor units of CP, whether concurrently or in subsequent transfusions within a week of the first CP transfusion. Due to low sample size in comparison to the HCA COVID-19 registry cohort, serology levels were not included as a covariate. Exploratory analyses were performed as described in results.

The day of the first CP transfusion was considered baseline (day 0) for the CP group. The comparison group was randomly assigned a pseudo-baseline prior to matching that reflected equal distribution of the time interval from admission to transfusion as the CP group (Supplemental Materials, Pseudo-Baseline).

### Outcomes Measures

#### All-cause in-hospital mortality

All patients were required to have a discharge date to be included in analyses. Patient vital status at time of discharge delineated censored from expired. Start time for all analyses was defined by baseline, which was the first CP transfusion date for the CP group and the assigned pseudo-baseline for the comparison group (Supplemental Materials, Pseudo-Baseline).

#### RTRM risk trajectory

Daily mortality risk scores were generated for each patient across the length of their hospitalization using probabilities from our COVID-19 Real-Time Risk Model (RTRM), which incorporates hundreds of structured medical record features such as clinico-demographics, comorbidities, laboratory values, secondary infections, complications, oxygenation details, and oxygen supplementation (22). When using the daily RTRM probabilities as a longitudinal outcome measure of progression and/or recovery, we also included baseline RTRM probability as a matching variable. Medications were purposely excluded from the RTRM model to be evaluated as covariates in comparative effectiveness studies such as this (Table S4).

### Statistics

#### Matching

All baseline patient characteristics were compared between the CP and comparison groups using t-tests for continuous variables and chi-squared tests for categorical variables. To mitigate bias resulting from non-randomized assignment of treatment, a Coarsened Exact Matching technique was used to match patients in the comparison group to the CP group (25). The matching for the main analyses was done based on patient age groupings, sex, race/ethnicity, significantly different comorbidities, calendar epoch, intubation, and baseline severity (WHO PS or RTRM probability in 0.10 increments) (26). Details on calendar epochs are provided in supplemental materials and Table S5. Co-variable imbalance pre- and post-matching was evaluated using the *L*_*1*_ statistic. Matching for all analyses were excellent with all post-matching *L*_*1*_ ≈ 0. Post-match datasets were generated specifically for each main and subgroup analysis as described in results (Figure 1).

The WHO PS is a modified 6-point scale, adapted from the WHO R&D Blueprint group to assess disease severity and measure clinical improvement in hospitalized patients (Table S6).(22, 27) Patients were assigned a daily WHO PS based on their most severe status that day.

#### Model fitting

All mortality analyses were examined for significant contributions of facility effect using a shared frailty model in comparison to a cox proportional hazards regression model (28). Additionally, the assumption of proportional hazards of a cox proportional hazards regression model was examined and, if violated, an accelerated failure time (AFT) model was implemented to examine consistency of effects (29).

We conducted a generalized linear mixed models (GLMM) approach for longitudinal data to examine trends in the daily RTRM probabilities over length-of-stay (RTRM trajectories) as a surrogate of progression/recovery. GLMM allowed for nesting of longitudinal observations under each patient included in the dataset. For RTRM trajectories, all nesting parameters were examined for significant contributions to the model. GLMM models included random intercepts for patient-level RTRM trajectories and facility nesting.

#### Subgroup analyses

Additional analyses were performed to examine the association of CP with mortality depending on a patient’s disease severity. Intubation is representative of severe disease. To perform subgroup analyses on intubated versus non-intubated patients, the cohorts were re-matched to exclude intubation as a matching variable so that the effect of intubation could be evaluated (Figure 1). Patients were considered intubated if they had a record of intubation prior to or on day of baseline for the intubation subgroup analysis.

Finally, we conducted exploratory analyses examining the association of days from admission to transfusion with mortality, specifically within the CP cohort. The initial model examined the days from admission to transfusion as a continuous variable in relationship with mortality outcomes. Additionally, we stratified the main RTRM sample into two different groups, 0-3 and 4-7 days, to examine the association between CP and mortality within a given transfusion window. We re-matched each of these cohort transfusion windows separately on baseline RTRM score, calendar epoch, intubation during hospitalization, sex, age, and race/ethnicity.

All analyses were performed using R v3.6.3 with the following packages: “cem”, “survival”, “coxme”, “lme4” (30). The pseudo-baseline assignment was conducted using an automated macro in SAS 9.4 (SAS Inc., Cary NC) (31).

### Human Participants and Study Approval

This study was supported by HCA Healthcare and conducted in accordance with US regulations, applicable ICH E6 international standards of Good Clinical Practice, and institutional research policies and procedures. This research was performed under a master retrospective protocol (MR 01) approved under expedited review by an external governing institutional review board (IntegReview/Advarra) and granted a waiver of informed consent.

## Data Availability

The data that support the findings of this study are available upon request from the corresponding author. The data are not publicly available due to privacy restrictions.

https://www.medrxiv.org/content/10.1101/2021.04.26.21256138v1

## Author Contributions

SAAE oversaw all aspects of the study including conceptualizing the study design and directing its implementation, as well as contributed to the development of the statistical analysis plan, interpretation of results, and manuscript writing and review. AJ contributed to the study design, developed the statistical analysis plan, performed all data cleanup and dataset construction, coded and implemented all analyses, interpreted results and contributed to manuscript writing and review. SAAE and AJ contributed equally. JSAR and MW provided clinical insight and contributed to results interpretation and manuscript preparation and review. CM contributed to EHR data curation and sourcing, data verification, and manuscript review and editing. PD performed preliminary data cleanup, analyses, pseudo-baseline assignments, and manuscript review and editing. DS contributed to RTRM modeling, results interpretation and manuscript preparation. LES provided oversight and contributed to overall study design, results interpretation and manuscript review. MC contributed to resource allocation, results interpretation and manuscript review. GF provided resource allocation, general oversight and contributed to results interpretation and manuscript review. HAB provided resource allocation, general oversight and contributed to results interpretation and manuscript review. CFL contributed to study conception and design, provided clinical insight and contributed to results interpretation and manuscript preparation and review.

## Acknowledgements

The authors wish to acknowledge Faisal Cheema, MD who contributed to operationalizing our HCA EAP CP program and provided clinical insight. Susan Garwood, MD also provided clinical insight. We would also like to thank Troy Gifford, Ryan Patrick, Chris Guthrie, Cecile Roman, Arielle Fisher, PhD, Jessica Correia, PhD, Daniel Luckett, PhD, Kathryn Hopkins-McGill, and Shaita Picard for their contributions to electronic data extraction, management, and curation for the HCA Healthcare COVID-19 registry. Thank you to Molly Altman and Andrew Stillwell for providing project management support. We would also like to acknowledge Mandelin Cooper, PharmD, Mitch Davis and Laura McLean in the Clinical Operations Group for their parallel COVID-19 work that provided insight into HCA Healthcare treatment recommendations for the date epochs as well as variable sourcing and mapping suggestions. Many thanks to Sybil Hyatt, Sally Mathews and the HRI Research Operations team as well as Michael Frost and the HRI Data Operations Team for their manual chart review efforts in confirming COVID-19 cases and identifying convalescent plasma cases early in the pandemic for the HCA Healthcare COVID-19 Registry. We are grateful for our Forward Pathology Solutions and Metro Infectious Disease Consultants colleagues in Kansas City, MO for their continued support of JR and MW collaborations. We would also like to acknowledge our blood-bank collaborators that provided insight and serology on donor plasma including Pampee Young, MD, PhD at the American Red Cross and Larry Dumont, MBA, PhD at Vitalant as well as our colleagues at Versiti. Finally, we would like to thank our collaborators at Mayo Clinic and Michigan State University, Rickey Carter, PhD and Nigel Paneth, MD, MPH, for their *post hoc* discussion of the findings in context with current methodologies and convalescent plasma literature.

## Supplemental Materials

### Supplemental Material Index

#### Supplemental Methods

Patient Encounters, Comparison Group Pseudo-Baseline Assignment, Epoch Intervals for Evolving Treatment Recommendations, Matching and Covariates

## Supplemental Methods

### Patient Encounters

For patients that had multiple hospital admissions within the date range of the data pull, we identified the longest encounter that was within the determined window of the first positive SARS-CoV-2 test and proceeded to link encounters, before and/or after, that were within 36 hours of that longest encounter’s admission and discharge times to create a continuum of care to adequately evaluate the disease course. For the CP group, the anchoring encounter included the first transfusion date of CP and before and after encounters within 36 hours were linked as already described.

### Comparison Group Pseudo-Baseline Assignment

The day of the first CP transfusion was considered baseline (day 0) for the CP group. The comparison group was automatically assigned a pseudo-baseline prior to matching that reflected equal distribution of the time interval from admission to transfusion as the CP group. Since the distribution of baseline dates was randomly assigned, there was a potential to assign a pseudo-baseline after a patient’s discharge date. To minimize this issue, we stratified the sample into different treatment epoch intervals before assigning the pseudo-baseline to the comparison group (Table S5). We removed any comparison patients that were discharged prior to their assigned pseudo-baseline (Figure 1)

### Epoch Intervals for Evolving Treatment Recommendations

The time interval from admission to CP transfusion decreased over time when investigating calendar dates. To account for this shift, we created date ranges that aligned with updates in HCA treatment recommendations which resulted in six epoch intervals which including March 2 to April 2, April 3 to April 29, April 30 to May 19, May 20 to July 5, July 6 to August 23, and August 24 to September 28. For an in-depth description of these clinico-demographic shifts, see Table S2. Additionally, we used epoch interval as a matching variable in our analyses. Since there were still major shifts in treatment approaches and hotspots for COVID19 cases within each epoch interval, we also included a covariate of a numerated calendar date for patient admission.

### Matching and Covariates

A variety of patient characteristics were captured at baseline that were considered for matching criteria. Demographic characteristics included age, sex, and race/ethnicity. Age ranges were grouped 18-44, 45-64, 65-74, 75-84, and over 84 (1). Baseline severity was matched on either the 6-point WHO PS, where only 2 to 5 were possible at baseline, or baseline RTRM probabilities in increments of 0.10. Additional clinical characteristics were included in matching such as preadmission comorbidities (as detailed below), admission epoch interval, and intubation during hospitalization. Due to missingness of smoking status (∼20%), we provided these data descriptively but excluded them from inferential analyses. Body mass index was found to be unreliably reported, especially for non-ambulatory patients, so it was excluded from all evaluations.

Covariates for models were considered for patient data that were related to the hospital stay. Treatment characteristics included pharmacy data: in-hospital administration of other medications inclusive of anticoagulants, statins/ACEi, steroids, other immunomodulators, antivirals, antibiotics, remdesivir, hydroxychloroquine (HCQ), tocilizumab, and azithromycin (Table S4), where ACEi is Angiotensin converting enzyme inhibitors. Azithromycin was considered a separate medication grouping from antibiotics because it was often used in conjunction with HCQ and/or for its additional anti-inflammatory properties. SARS-CoV-2 monoclonal antibodies were not authorized (EUA) for use in hospitalized patients prior to our data pull so no instances of monoclonal infusion occurred in our cohort (2). Medications were computed as indicators of usage over the length of hospitalization. Secondary infections were considered such as bacterial pneumonia, sepsis, and severe sepsis but were included in the RTRM calculations and, thus, were not included in the RTRM-matched models. However, secondary infections were included as covariates in the WHO PS-matched models. Although epoch intervals were included in the matching criteria, we included numerated calendar date as a covariate to account for the fluctuations within the epoch intervals. Finally, since all inferential analyses started with day 0 at transfusion date/pseudo-baseline, the number of days from admission to baseline was also included in the models as a covariate.

Comorbidities included in the RTRM were based on the Charlson Comorbidity Index and included diabetes with chronic complications, diabetes without chronic complications, all-inclusive diabetes, hypertension, chronic ischemic heart disease, congestive heart failure, renal disease (mild or moderate), renal disease (severe), asthma or reactive airway disease, chronic pulmonary disease (excluding asthma), HIV infection, cancer (including solid tumors and blood cancers but excluding non-melanoma skin cancer), and recently diagnosed cancer within two years of admission date (3, 4). For the current analyses, we also accounted for both organ-specific and systemic autoimmune diseases (5).

### Analytic Software

All analyses were performed using R v3.6.3 with the following packages: “cem”, “survival”, “coxme”, “lme4” (6). The pseudo-baseline assignment was conducted using an automated macro in SAS 9.4 (SAS Inc., Cary NC) (7).

---

**Figure S1.**
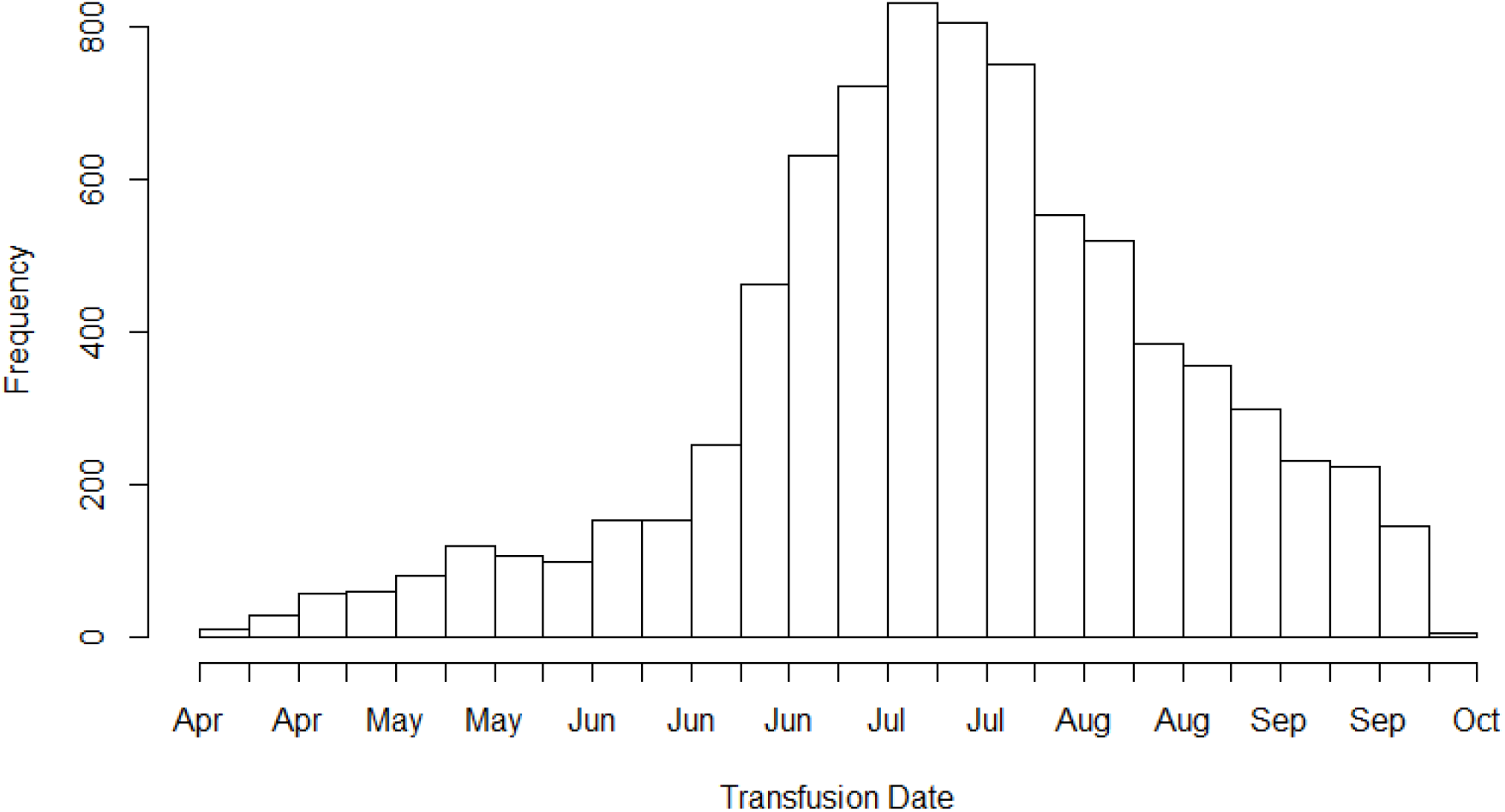
Distribution Frequencies of CP Transfusions by Calendar Date.

**Figure S2.**
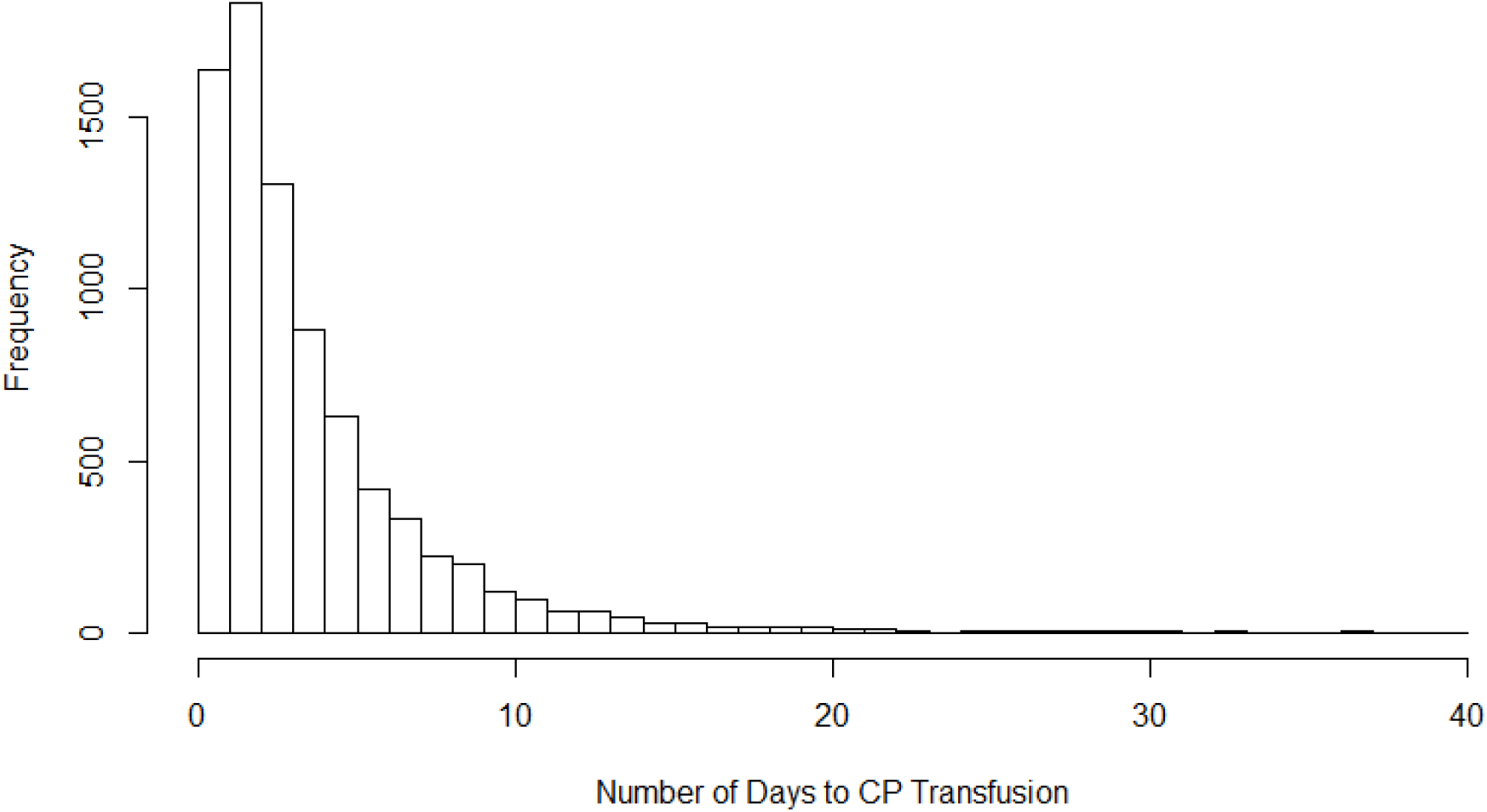
Distribution Frequencies of Days from Admission to Transfusion with CP.

## Supplemental Tables

**Table S1.**
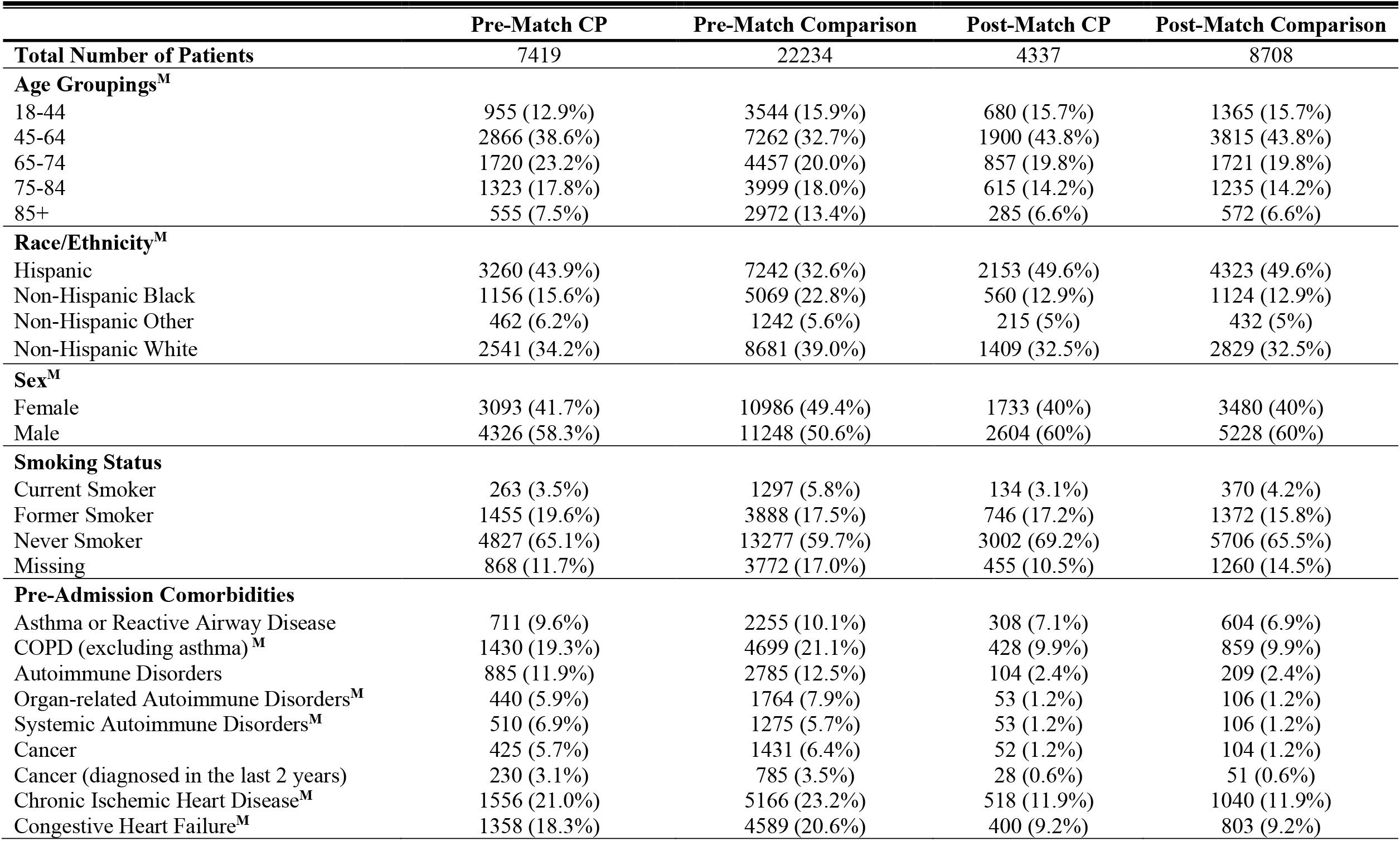
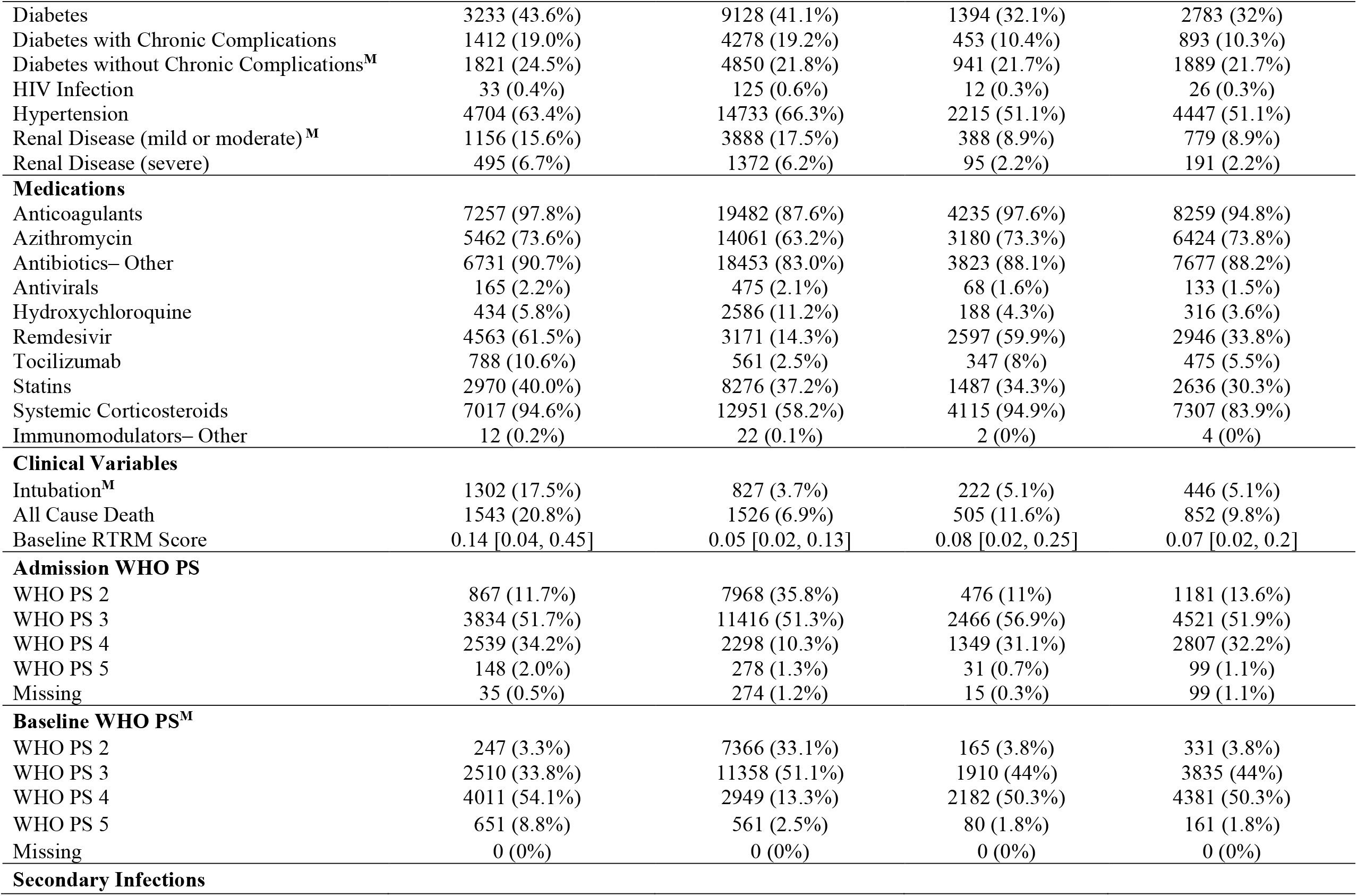
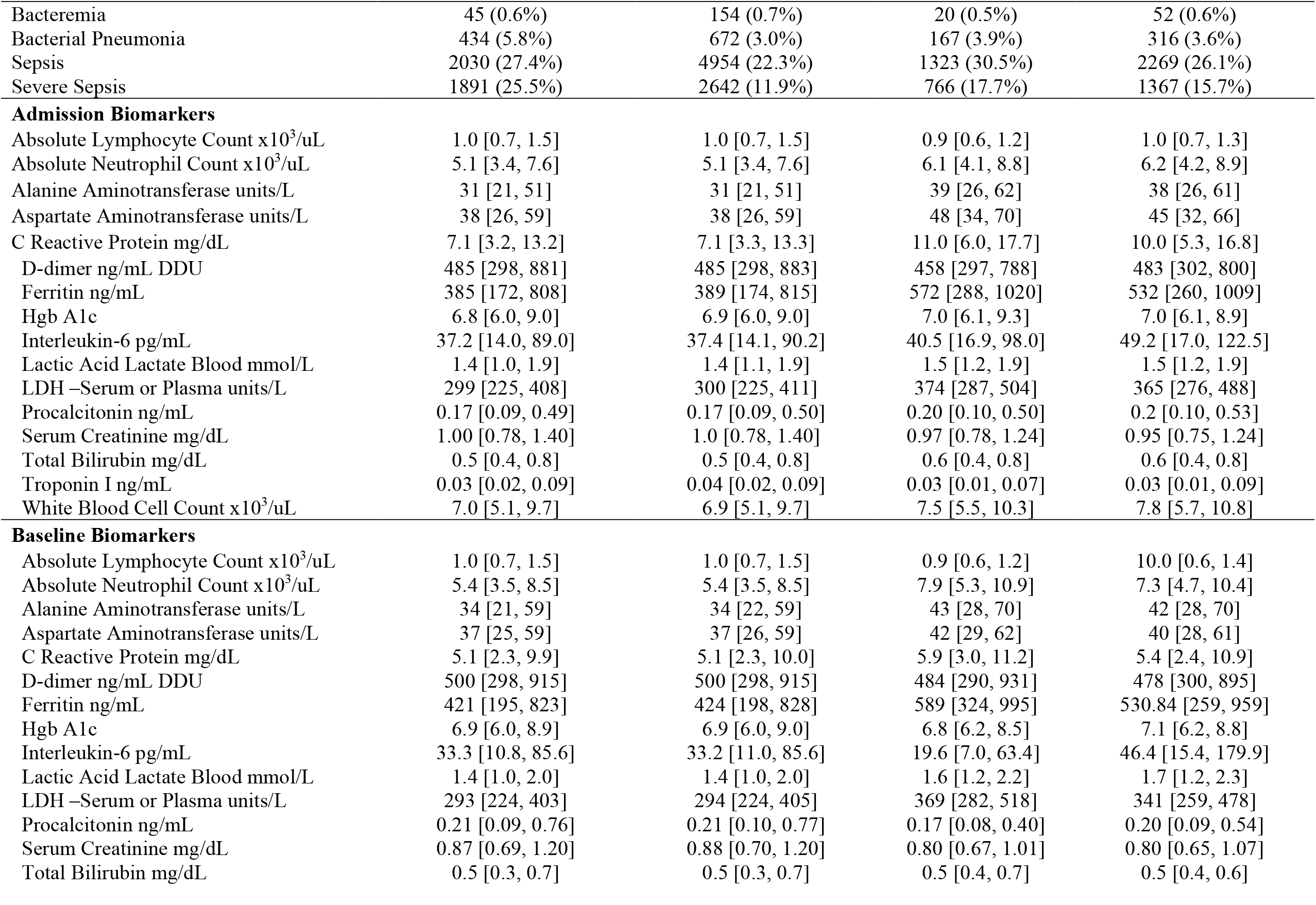
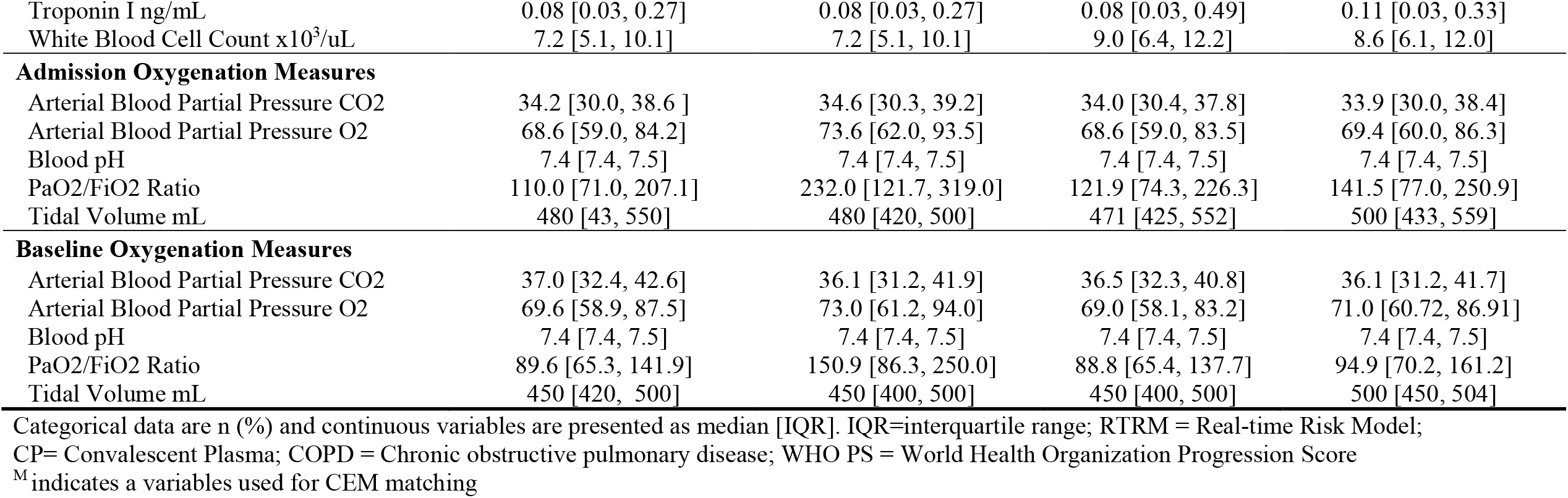
Pre- and Post-Match Table: WHO PS-Matched Model. Frequencies and distributions of characteristics, comorbid conditions, and admission and baseline laboratory results pre- and post-matching for both CP and comparison groups.

**Table S2.** Variable Description of Epoch Intervals. Frequencies and distributions of characteristics, comorbid conditions, and admission and baseline laboratory results over key calendar epochs for patients hospitalized with COVID-19.

|  | **All Patients** | **EPOCH 1** | **EPOCH 2** | **EPOCH 3** | **EPOCH 4** | **EPOCH 5** | **EPOCH 6** |
| --- | --- | --- | --- | --- | --- | --- | --- |
| **Total Number of Patients, N** | 33987 | 1380 | 2668 | 1585 | 8527 | 15753 | 4074 |
| **Number of CP Patients, N** | 8034 | 10 | 158 | 260 | 2173 | 4281 | 1152 |
| **Age Groupings** |  |  |  |  |  |  |  |
| 18-44 | 5734 (16.9%) | 215 (15.6%) | 370 (13.9%) | 277 (17.5%) | 1821 (21.4%) | 2435 (15.5%) | 616 (15.1%) |
| 45-64 | 11481 (33.8%) | 523 (37.9%) | 878 (32.9%) | 526 (33.2%) | 3141 (36.8%) | 5143 (32.6%) | 1270 (31.2%) |
| 65-74 | 6942 (20.4%) | 298 (21.6%) | 545 (20.4%) | 292 (18.4%) | 1551 (18.2%) | 3378 (21.4%) | 878 (21.6%) |
| 75-84 | 5961 (17.5%) | 208 (15.1%) | 477 (17.9%) | 272 (17.2%) | 1229 (14.4%) | 2956 (18.8%) | 819 (20.1%) |
| 85+ | 3862 (11.4%) | 135 (9.8%) | 398 (14.9%) | 217 (13.7%) | 783 (9.2%) | 1838 (11.7%) | 491 (12.1%) |
| **Race/Ethnicity** |  |  |  |  |  |  |  |
| Hispanic | 11040 (32.5%) | 290 (21%) | 664 (24.9%) | 483 (30.5%) | 3510 (41.2%) | 5121 (32.5%) | 972 (23.9%) |
| Non-Hispanic Black | 6479 (19.1%) | 389 (28.2%) | 600 (22.5%) | 324 (20.4%) | 1522 (17.8%) | 2985 (18.9%) | 659 (16.2%) |
| Non-Hispanic Other | 1810 (5.3%) | 97 (7%) | 200 (7.5%) | 101 (6.4%) | 463 (5.4%) | 763 (4.8%) | 186 (4.6%) |
| Non-Hispanic White | 11823 (34.8%) | 481 (34.9%) | 1011 (37.9%) | 534 (33.7%) | 2375 (27.9%) | 5585 (35.5%) | 1837 (45.1%) |
| **Sex** |  |  |  |  |  |  |  |
| Female | 16430 (48.3%) | 645 (46.7%) | 1298 (48.7%) | 755 (47.6%) | 4052 (47.5%) | 7635 (48.5%) | 2045 (50.2%) |
| Male | 17547 (51.6%) | 734 (53.2%) | 1370 (51.3%) | 829 (52.3%) | 4473 (52.5%) | 8113 (51.5%) | 2028 (49.8%) |
| Unknown | 3 (0%) | 0 (0%) | 0 (0%) | 0 (0%) | 0 (0%) | 2 (0%) | 1 (0%) |
| **Pre-Admission Comorbidities** |  |  |  |  |  |  |  |
| Asthma or Reactive Airway Disease | 3251 (9.6%) | 165 (12%) | 283 (10.6%) | 139 (8.8%) | 802 (9.4%) | 1468 (9.3%) | 394 (9.7%) |
| COPD (Excluding Asthma) | 6615 (19.5%) | 260 (18.8%) | 542 (20.3%) | 325 (20.5%) | 1410 (16.5%) | 3240 (20.6%) | 838 (20.6%) |
| Autoimmune Disorders total | 4008 (11.8%) | 154 (11.2%) | 363 (13.6%) | 204 (12.9%) | 905 (10.6%) | 1865 (11.8%) | 517 (12.7%) |
| Organ-related Autoimmune Disorders | 2390 (7%) | 91 (6.6%) | 207 (7.8%) | 131 (8.3%) | 516 (6.1%) | 1156 (7.3%) | 289 (7.1%) |
| Systemic Autoimmune Disorders | 1956 (5.8%) | 78 (5.7%) | 182 (6.8%) | 95 (6%) | 469 (5.5%) | 865 (5.5%) | 267 (6.6%) |
| Cancer | 2021 (5.9%) | 83 (6%) | 182 (6.8%) | 76 (4.8%) | 441 (5.2%) | 969 (6.2%) | 270 (6.6%) |
| Cancer (diagnosed in the last 2 years) | 1132 (3.3%) | 43 (3.1%) | 106 (4%) | 41 (2.6%) | 254 (3%) | 538 (3.4%) | 150 (3.7%) |
| Chronic Ischemic Heart Disease | 7257 (21.4%) | 269 (19.5%) | 599 (22.5%) | 354 (22.3%) | 1512 (17.7%) | 3572 (22.7%) | 951 (23.3%) |
| Congestive Heart Failure | 6481 (19.1%) | 245 (17.8%) | 616 (23.1%) | 309 (19.5%) | 1367 (16%) | 3101 (19.7%) | 843 (20.7%) |
| Diabetes total | 13545 (39.9%) | 539 (39.1%) | 1128 (42.3%) | 608 (38.4%) | 3235 (37.9%) | 6507 (41.3%) | 1528 (37.5%) |
| Diabetes with Chronic Complications | 6154 (18.1%) | 232 (16.8%) | 517 (19.4%) | 300 (18.9%) | 1312 (15.4%) | 3056 (19.4%) | 737 (18.1%) |
| Diabetes without Chronic Complications | 7391 (21.7%) | 307 (22.2%) | 611 (22.9%) | 308 (19.4%) | 1923 (22.6%) | 3451 (21.9%) | 791 (19.4%) |
| HIV Infection | 171 (0.5%) | 8 (0.6%) | 16 (0.6%) | 11 (0.7%) | 56 (0.7%) | 73 (0.5%) | 7 (0.2%) |
| Hypertension | 21368 (62.9%) | 951 (68.9%) | 1825 (68.4%) | 996 (62.8%) | 4977 (58.4%) | 10059 (63.9%) | 2560 (62.8%) |
| Renal Disease (mild or moderate) | 5497 (16.2%) | 230 (16.7%) | 486 (18.2%) | 271 (17.1%) | 1147 (13.5%) | 2666 (16.9%) | 697 (17.1%) |
| Renal Disease (severe) | 2021 (5.9%) | 77 (5.6%) | 187 (7%) | 117 (7.4%) | 440 (5.2%) | 991 (6.3%) | 209 (5.1%) |
| **Medications** |  |  |  |  |  |  |  |
| Anticoagulants | 29233 (86%) | 1020 (73.9%) | 2161 (81%) | 1357 (85.6%) | 7569 (88.8%) | 13835 (87.8%) | 3291 (80.8%) |
| Azithromycin | 21167 (62.3%) | 1140 (82.6%) | 1846 (69.2%) | 976 (61.6%) | 5475 (64.2%) | 9584 (60.8%) | 2146 (52.7%) |
| Antibiotics – Other | 27507 (80.9%) | 1196 (86.7%) | 2185 (81.9%) | 1301 (82.1%) | 7100 (83.3%) | 12703 (80.6%) | 3022 (74.2%) |
| Antivirals | 697 (2.1%) | 134 (9.7%) | 91 (3.4%) | 39 (2.5%) | 152 (1.8%) | 241 (1.5%) | 40 (1%) |
| Hydroxychloroquine | 3282 (9.7%) | 793 (57.5%) | 1435 (53.8%) | 413 (26.1%) | 342 (4.0%) | 265 (1.7%) | 34 (0.8%) |
| Remdesivir | 8303 (24.4%) | 11 (0.8%) | 41 (1.5%) | 136 (8.6%) | 2510 (29.4%) | 4389 (27.9%) | 1216 (29.8%) |
| Tocilizumab | 1466 (4.3%) | 26 (1.9%) | 179 (6.7%) | 131 (8.3%) | 574 (6.7%) | 549 (3.5%) | 7 (0.2%) |
| Statins/ACE Inhibitors (ACEi) | 12100 (35.6%) | 476 (34.5%) | 1033 (38.7%) | 576 (36.3%) | 2746 (32.2%) | 5771 (36.6%) | 1498 (36.8%) |
| Steroids | 21690 (63.8%) | 290 (21%) | 699 (26.2%) | 482 (30.4%) | 5437 (63.8%) | 11967 (76%) | 2815 (69.1%) |
| Immunomodulators – Other | 37 (0.1%) | 2 (0.1%) | 4 (0.1%) | 2 (0.1%) | 12 (0.1%) | 13 (0.1%) | 4 (0.1%) |
| **Clinical Variables** |  |  |  |  |  |  |  |
| Days to transfusion (CP only), $M$ ($SD$) | 4.0 (3.7) | 20.9 (4.9) | 8.8 (6.4) | 4.4 (3.4) | 4.4 (4.0) | 4.0 (3.4) | 2.4 (2.1) |
| Length of Stay, $M$ ($SD$) | 9.5 (9.2) | 10.6 (11.7) | 11.4 (11.9) | 11.3 (10.5) | 10.2 (10.2) | 9.3 (8.4) | 6.8 (5.2) |
| Intubation | 2277 (6.7%) | 156 (11.3%) | 203 (7.6%) | 104 (6.6%) | 608 (7.1%) | 1053 (6.7%) | 153 (3.8%) |
| All Cause Death | 3431 (10.1%) | 158 (11.4%) | 310 (11.6%) | 134 (8.5%) | 827 (9.7%) | 1692 (10.7%) | 310 (7.6%) |
| **Admission WHO PS** |  |  |  |  |  |  |  |
| WHO PS 2 | 9572 (28.2%) | 413 (29.9%) | 741 (27.8%) | 479 (30.2%) | 2482 (29.1%) | 4262 (27.1%) | 1195 (29.3%) |
| WHO PS 3 | 16489 (48.5%) | 762 (55.2%) | 1454 (54.5%) | 796 (50.2%) | 4106 (48.2%) | 7498 (47.6%) | 1873 (46%) |
| WHO PS 4 | 5237 (15.4%) | 87 (6.3%) | 288 (10.8%) | 191 (12.1%) | 1318 (15.5%) | 2787 (17.7%) | 566 (13.9%) |
| WHO PS 5 | 539 (1.6%) | 57 (4.1%) | 56 (2.1%) | 21 (1.3%) | 127 (1.5%) | 228 (1.4%) | 50 (1.2%) |
| Missing | 2150 (6.3%) | 61 (4.4%) | 129 (4.8%) | 98 (6.2%) | 494 (5.8%) | 978 (6.2%) | 390 (9.6%) |
| **Baseline WHO PS** |  |  |  |  |  |  |  |
| WHO PS 2 | 8108 (23.9%) | 288 (20.9%) | 669 (25.1%) | 403 (25.4%) | 2046 (24%) | 3596 (22.8%) | 1106 (27.1%) |
| WHO PS 3 | 14853 (43.7%) | 695 (50.4%) | 1288 (48.3%) | 712 (44.9%) | 3637 (42.7%) | 6772 (43%) | 1749 (42.9%) |
| WHO PS 4 | 7489 (22%) | 148 (10.7%) | 390 (14.6%) | 302 (19.1%) | 2032 (23.8%) | 3862 (24.5%) | 755 (18.5%) |
| WHO PS 5 | 1331 (3.9%) | 172 (12.5%) | 177 (6.6%) | 57 (3.6%) | 320 (3.8%) | 540 (3.4%) | 65 (1.6%) |
| Missing | 2206 (6.5%) | 77 (5.6%) | 144 (5.4%) | 111 (7.0%) | 492 (5.8%) | 983 (6.2%) | 399 (9.8%) |
| **Secondary Infections** |  |  |  |  |  |  |  |
| Bacteremia | 221 (0.7%) | 7 (0.5%) | 15 (0.6%) | 12 (0.8%) | 44 (0.5%) | 115 (0.7%) | 28 (0.7%) |
| Bacterial Pneumonia | 1218 (3.6%) | 87 (6.3%) | 115 (4.3%) | 61 (3.8%) | 294 (3.4%) | 538 (3.4%) | 123 (3%) |
| Sepsis | 7756 (22.8%) | 321 (23.3%) | 637 (23.9%) | 393 (24.8%) | 2041 (23.9%) | 3515 (22.3%) | 849 (20.8%) |
| Severe Sepsis | 5045 (14.8%) | 255 (18.5%) | 501 (18.8%) | 243 (15.3%) | 1226 (14.4%) | 2288 (14.5%) | 532 (13.1%) |

Data are n (%) unless otherwise indicated. Epochs were all in 2020 and were defined as follows based on changes in treatment recommendations: Epoch 1, March 1 to April 2; Epoch 2, April 3 to April 29; Epoch 3, April 30 to May 19; Epoch 4, May 20 to July 5; Epoch 5, July 6 to August 23; Epoch 6, August 24 to September 28. CP= Convalescent Plasma; COPD = Chronic obstructive pulmonary disease; WHO PS = World Health Organization Progression Score

**Table S3.** Effects and Interactions of Serology in the CP Cohort. Results of the exploratory analyses on the relationship of serology levels with all-cause, in-hospital mortality in the CP cohort only where serology data was provided (N = 1944). In a Cox regression models, Ortho Vitros S/Co Total anti-SARS-CoV-2 serology levels were treated as continuous and ordinal by 20^th^ and 80^th^ percentiles for low, medium and high trend assessment. Further analyses looked at the interaction of serology level with days from admission to transfusion (N = 1939). Simple slope analyses were conducted for the decomposition of the significant interaction two separate ways: to examine the impact of high serology versus low serology across number of days from admission to transfusion, and to examine the impact of 0-3 days to transfusions versus 4+ days to transfusion across level of serology.

|  | HR | LLCI | ULCI | *p*-value |
| --- | --- | --- | --- | --- |
| **Continuous S/Co Levels** |  |  |  |  |
| Serology | 0.999 | 0.999 | 1.000 | 0.084 |
| **Categorical S/Co Levels** |  |  |  |  |
| Low (below 20<sup>th</sup> percentile)<sup>R</sup> | -- | -- | -- | -- |
| Medium (20<sup>th</sup> – 80<sup>th</sup> percentile) | 0.864 | 0.657 | 1.137 | 0.297 |
| High (above 80<sup>th</sup> percentile) | 0.777 | 0.548 | 1.101 | 0.155 |
| **S/Co Levels by Days to Transfusion** |  |  |  |  |
| Serology | **0.998** | 0.997 | 0.999 | **0.013** |
| Days to Transfusion | 1.036 | 1.002 | 1.071 | **0.037** |
| Serology * Days to Transfusion | 1.0002 | 1.0000 | 1.0004 | **0.044** |
| **Effects from Simple Slopes by Serology** |  |  |  |  |
| Low Serology: Days to Transfusion | 1.043 | 1.014 | 1.073 | **0.003** |
| High Serology: Days to Transfusion | 1.097 | 1.056 | 1.139 | **<0.001** |
| **Effects from Simple Slopes by Days** |  |  |  |  |
| 0-3 Days: Serology | **0.998** | 0.997 | 0.999 | **0.029** |
| 4+ Days: Serology | 1.000 | 0.999 | 1.001 | 0.736 |

For cases where patients received more than one CP transfusion within 1 week of their first CP transfusion, the serologic levels were averaged to create one serologic level per patient.
<sup>R</sup> represents the reference variable for the categorical S/Co serology levels. N = 1944
Five patients with erroneous admission dates were removed for all results that included Days to Transfusion. N = 1939
Simple slope analyses were conducted for the decomposition of the significant interaction; the transformed scores were centered by mean ± SD ( $178.21 \pm 138.49$ , N=1939) to examine the impact of high serology versus low serology across number of days from admission to transfusion as separate models.

**Table S4.** Medication List. Description of medications that were included into each medication category.

| **Medication Categories** | **List of Included Medications** |
| --- | --- |
| Anticoagulants | anticoagulant, acd, argatroban, arixtra, coumadin, eliquis, enoxaparin, heparin, lovenox, fondaparinux, warfarin, apixaban |
| Tocilizumab | actemra, tocilizumab |
| Azithromycin | azithromycin, zithromax, zmax |
| Statins/ACE Inhibitors (ACEi) | Statins: statin, crestor, lipitor, pravachol, pravastatin, rosuvastatin, zocor, atorvastatin, simvastatin<br>ACEi: ACE, ramipril, capoten, prinivil, vasotec, zestril, captopril, lisinopril, enalapril |
| Steroids | steroid, corticosteroid, cortef, hydrocortisone, decadron, dexamethasone, deltasone, prednisone, deltisone, solu-cortef, hydrocortisone, solu-medrol, methylprednisolone |
| Other Immunomodulators | imuran, myfortic, azathioprine, mycophenolic |
| Hydroxychloroquine | plaquenil, hydroxychloroquine |
| Remdesivir | remdesivir, veklury |
| Antivirals | antiviral, acyclovir, zovirax, abacavir, kivexa, ziagen, epzicom, trizivir, triumeq, tamiflu, oseltamivir, nrti, epivir, lamivudine |
| Other Antibiotics | amoxicillin, amoxil, augmentin, moxatag, ampicillin, principen, unasyn, oxacillin, penicillin, pfizerpen, bicillin, ancef, cefazolin, cefepime, maxipime, ceftin, cefuroxime, cefdinir, omnicef, fortaz, ceftazidime, keflex, cephalixin, kefzol, rocephin, ceftriaxone, zerbaxa, ceftolozane, cephalosporin, azactam, aztreona, monobactam, bactrim, trimethoprim, sulfonamide, cipro, ciprofloxacin, levaquin, levofloxacin, fluoroquinolone, cleocin, clindamycin, lincosamide, cubicin, daptomycin, flagyl, metronidazole, invanz, ertapenem, merrem, meropenem, carbapenem, macrobid, nitrofurantoin, macrodantin, nebcin, aminoglycoside, sulfamethoxazole, gantanol, sulfatrim, zotrim, vancocin, vancomycin, firvanq, xifaxan, rifaximin, rifamycin, rimactane, aemcolo, zosyn, piperacillin, zyvox, linezolid |

**Table S5.** Calendar Epoch Intervals. Definitions of six calendar epochs used to account for changes in HCA treatment recommendations for patients hospitalized with COVID-19 over time in 2020.

| Epoch | Calendar Definitions | Summary of Treatment Recommendations |
| --- | --- | --- |
| Epoch 1 | March 2 – April 2, 2020 | Consider only hydroxychloroquine without azithromycin<br>Consider remdesivir<br>Avoid steroids |
| Epoch 2 | April 3 – April 29, 2020 | Consider hydroxychloroquine with azithromycin<br>Consider tocilizumab<br>Consider convalescent plasma |
| Epoch 3 | April 30 – May 19, 2020 | Consider only hydroxychloroquine, no azithromycin<br>Consider low-dose, short-course steroids in later pulmonary phase |
| Epoch 4 | May 20 – July 5, 2020 | Do not consider hydroxychloroquine |
| Epoch 5 | July 6 – August 23, 2020 | Steroids recommended for all patients requiring oxygen supplementation |
| Epoch 6 | August 24 – September 28, 2020 | Consider convalescent plasma for patients with lesser severity illness* |

\*Shift in treatment recommendation for convalescent plasma from patient with more severe disease to patients with less severe disease based on the shift from the expanded access program (EAP) requirements to the emergency use authorization (EUA).(4)

**Table S6.**
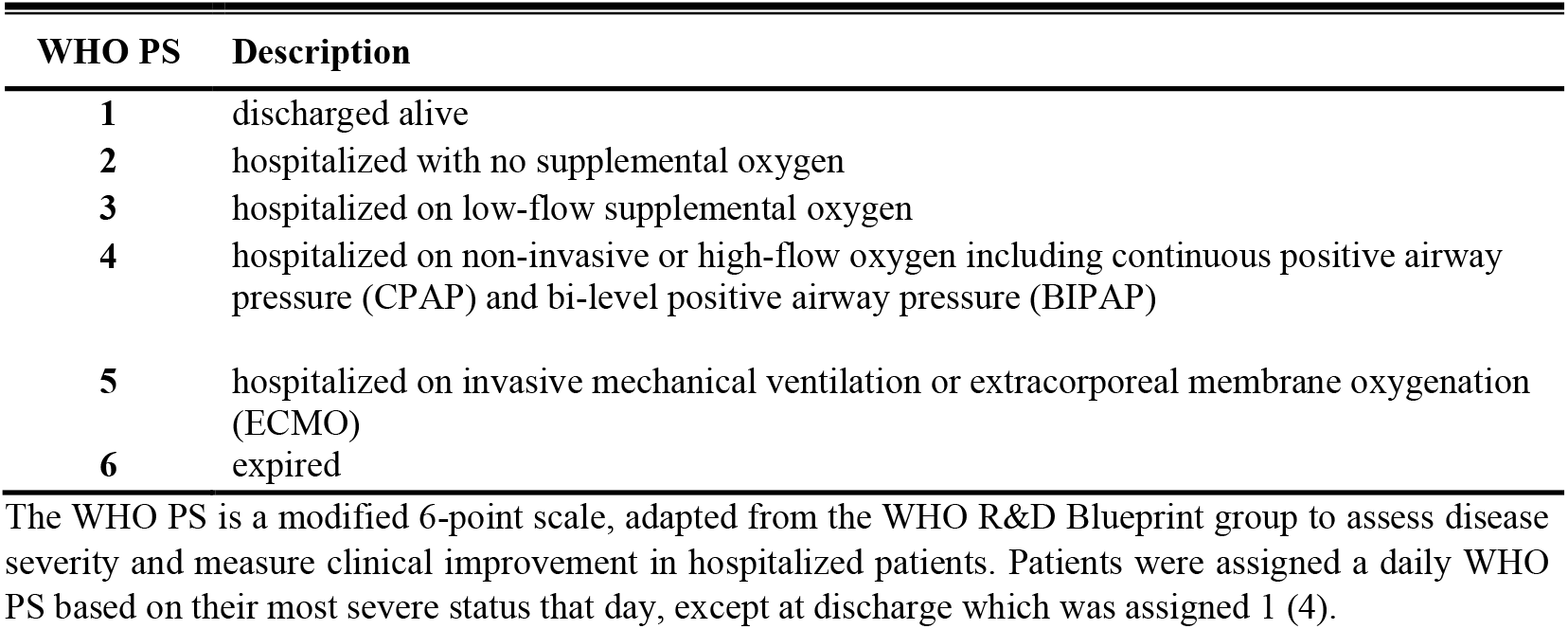
WHO Progression Score Categories. List defining the modified 6-point WHO progression score categories (WHO PS) used to match for baseline disease severity.

| WHO PS | Description |
| --- | --- |
| 1 | discharged alive |
| 2 | hospitalized with no supplemental oxygen |
| 3 | hospitalized on low-flow supplemental oxygen |
| 4 | hospitalized on non-invasive or high-flow oxygen including continuous positive airway pressure (CPAP) and bi-level positive airway pressure (BIPAP) |
| 5 | hospitalized on invasive mechanical ventilation or extracorporeal membrane oxygenation (ECMO) |
| 6 | expired |

The WHO PS is a modified 6-point scale, adapted from the WHO R&D Blueprint group to assess disease severity and measure clinical improvement in hospitalized patients. Patients were assigned a daily WHO PS based on their most severe status that day, except at discharge which was assigned 1 (4).

